# Outcomes and phenotypic expression of rare variants in hypertrophic cardiomyopathy genes amongst UK Biobank participants

**DOI:** 10.1101/2021.01.21.21249470

**Authors:** Antonio de Marvao, Kathryn A McGurk, Sean L Zheng, Marjola Thanaj, Wenjia Bai, Jinming Duan, Carlo Biffi, Francesco Mazzarotto, Ben Statton, Timothy JW Dawes, Nicolò Savioli, Brian P Halliday, Xiao Xu, Rachel J Buchan, A John Baksi, Marina Quinlan, Paweł Tokarczuk, Upasana Tayal, Catherine Francis, Nicola Whiffin, Pantazis I Theotokis, Xiaolei Zhang, Mikyung Jang, Alaine Berry, Antonis Pantazis, Paul JR Barton, Daniel Rueckert, Sanjay K Prasad, Roddy Walsh, Carolyn Y Ho, Stuart A Cook, James S Ware, Declan P O’Regan

## Abstract

**Background:** Hypertrophic cardiomyopathy (HCM) is caused by rare variants in sarcomere-encoding genes, but little is known about the clinical significance of these variants in the general population.

**Methods:** We compared outcomes and cardiovascular phenotypes in UK Biobank participants with whole exome sequencing stratified by sarcomere-encoding variant status.

**Results:** The prevalence of rare variants (allele frequency <0.00004) in HCM-associated sarcomere-encoding genes in 200,584 participants was 2.9% (n=5,727; 1 in 35), of which 0.24% (n=474, 1 in 423) were pathogenic or likely pathogenic variants (SARC-P/LP). SARC-P/LP variants were associated with increased risk of death or major adverse cardiac events compared to controls (HR 1.68, 95% CI 1.37-2.06, p<0.001), mainly due to heart failure (HR 4.40, 95% CI 3.22-6.02, p<0.001) and arrhythmia (HR 1.55, 95% CI 1.18-2.03, p=0.002). In 21,322 participants with cardiac magnetic resonance imaging, SARC-P/LP were associated with increased left ventricular maximum wall thickness (10.9±2.7 vs 9.4±1.6 mm, p<0.001) and concentric remodelling (mass/volume ratio: 0.63±0.12 vs 0.58±0.09 g/mL, p<0.001), but hypertrophy (≥13mm) was only present in 16% (n=7/43, 95% CI 7-31%). Other rare sarcomere-encoding variants had a weak effect on wall thickness (9.5±1.7 vs 9.4±1.6 mm, p=0.002) with no combined excess cardiovascular risk (HR 1.00 95% CI 0.92-1.08, p=0.9).

**Conclusions:** In the general population, SARC-P/LP variants have low aggregate penetrance for overt HCM but are associated with an increased risk of adverse cardiovascular outcomes and a sub-clinical cardiomyopathic phenotype. In contrast, rare sarcomeric variants that do not meet criteria to be classified as P/LP appear to have minimal clinical impact.

## Introduction

Hypertrophic cardiomyopathy (HCM) is the most common genetic cardiomyopathy and is characterised by clinical and genetic heterogeneity, incomplete and age-dependent penetrance, and variable expressivity.^1^ Most individuals with HCM have a normal life expectancy but an important subset is at increased risk of adverse outcomes such as heart failure, atrial fibrillation, stroke or sudden cardiac death, with the cumulative incidence of morbidity and mortality reaching ∼77% by 60 years of age.^2^ The prevalence of overt disease in young adults has been reported as 1 in 500,^3^although wider use of genetic testing indicates that a significantly higher number individuals, up to 1 in 200, are carriers of pathogenic / likely pathogenic variants.^4^

Genetic variation in eight genes encoding sarcomeric components, principally *MYH7* and *MYBPC3*, account for the majority of genetically explained HCM.^5^Genetic testing identifies such variants in approximately 50% of cases, enabling cascade screening of first-degree relatives, and identification of at-risk variant carriers.^6^In those diagnosed with HCM, the presence of pathogenic sarcomeric variants is a strong predictor of worse outcomes.^2^However, little is known about the prevalence and phenotypic expression of sarcomeric variants outside of families with penetrant disease. Moreover, cardiovascular outcomes in sarcomere variant carriers in the general population have not yet been characterised and it is unclear if adverse clinical events occur only in individuals that express an HCM phenotype. Determining the significance of rare HCM-associated variants or of incidental unexplained hypertrophy remains a key challenge of contemporary clinical genetics and cardiovascular medicine.^7^

To determine the population prevalence of HCM-associated sarcomeric variants, characterise their phenotypic manifestations, estimate penetrance, and identify associations between sarcomeric variants and clinical outcomes, we performed an observational study of 218,813 adults in the UK Biobank (UKBB), of whom 200,584 have whole exome sequencing (WES).

## Methods

### Study Cohorts

The UKBB recruited 500,000 participants aged 40–69 years across the United Kingdom between 2006 and 2010 (National Research Ethics Service - 11/NW/0382).^8^A statement on research ethics approval for this biobank is available online (https://www.ukbiobank.ac.uk/learn-more-about-uk-biobank/about-us/ethics). In each case written informed consent was provided. Approval was received to use the data in the present work under terms of access approval number 40616. The results are reported in accordance with the Strengthening the Reporting of Observational Studies in Epidemiology (STROBE) guidelines^9^and checklist provided in **Table S1**.

### Cardiac phenotyping using machine learning

Participants in the imaging sub-study underwent cardiac magnetic resonance imaging (CMR) at 1.5T according to standardised published protocols.^10^ Segmentation of the cine images was performed using a deep learning neural network algorithm developed in-house and optimised on the UKBB cohort. The performance of image annotation using this algorithm is equivalent to a consensus of expert human readers and achieves sub-pixel accuracy for cardiac segmentation.^11^ Myocardial wall thickness (WT) was measured along radial line segments connecting the endocardial and epicardial surfaces perpendicular to the myocardial centreline and excluding trabeculae (**Figure 1**) – an approach that also exceeds the performance of human experts.^12^ Chamber volumes and mass were calculated from the segmentations according to standard post-processing guidelines.^13^ Myocardial strain analysis was performed using non-rigid free-from deformation image registration.^14,15^ Trabecular traits were quantified using fractal dimension (FD) analysis where a higher value indicates more complex trabeculation.^16^ Parametric three-dimensional analysis of the geometry of the left ventricle was performed to map regional patterns of remodelling and quantify the association with genetic and environmental predictors.^14,17-19^ Further details on phenotyping are given in the Supplementary Appendix.

**Figure 1.**
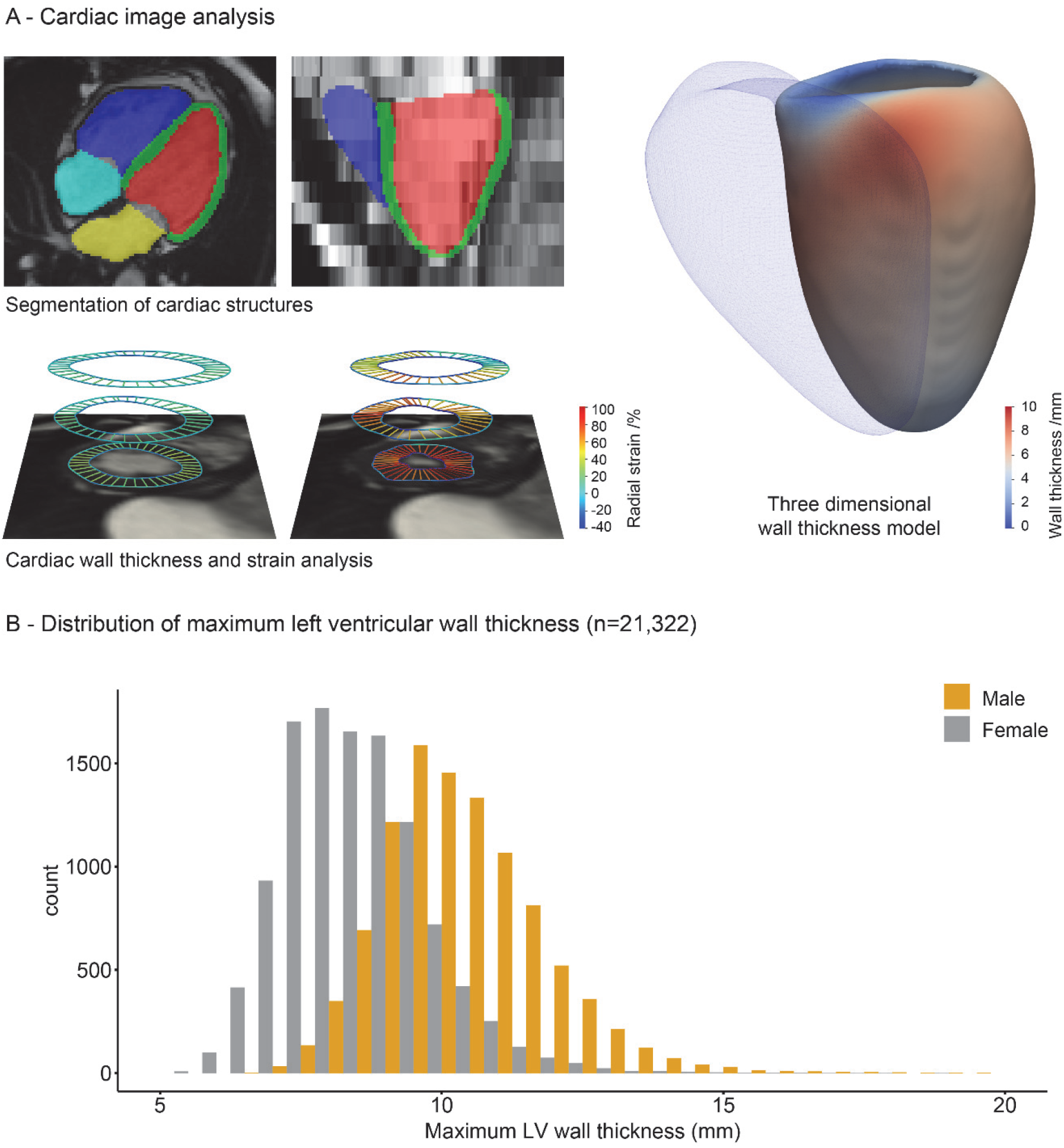
A) Computer vision techniques were used to automatically segment the four chambers of the heart from cardiac magnetic resonance imaging (right atrium: light blue; right ventricle: dark blue; left atrium: yellow; left ventricle: red; left ventricular myocardium: green). Cardiac motion analysis was used to derive strain and strain rates (diastole and systole shown with colours representing radial strain). Regional analysis of left ventricular wall thickness was performed using three-dimensional modelling of these segmentations aligned to a common reference space. Here, mean wall thickness for 21,322 UK Biobank participants is mapped onto the surface of the left ventricle with the position of the right ventricle shown as a mesh. B). Histogram of maximum left ventricular wall thickness by sex in UK Biobank.

### Sequencing and variant categorisation pipeline

UKBB participants underwent WES as previously described.^20^ They were divided into three genetic strata. Individuals were classified as genotype negative (**SARC-NEG**) if they had no rare protein-altering genetic variation (minor allele frequency (MAF) <0.001 in UKBB or the Genome Aggregation Database^21^) in any of 25 genes that may cause or mimic HCM. These 25 genes represent an inclusive list of 8 definitive-evidence sarcomere-encoding genes, 3 moderate-evidence sarcomere-encoding genes, and 14 genes associated with syndromic phenotypes that can include left ventricular (LV) hypertrophy.^22^This SARC-NEG group was compared to carriers of rare variants in 8 sarcomere-encoding genes definitively associated with HCM (*MYBPC3, MYH7, MYL2, MYL3, TNNI3, TNNT2, TPM1, ACTC1*).^22^Analysis was restricted to robustly associated variant classes for each gene, and to variants sufficiently rare to cause penetrant disease (filtering allele frequency (FAF) <0.00004).^23^

Variants were classified as pathogenic/likely pathogenic (**SARC-P/LP**) if previously observed in an individual with HCM and with at least 2-star evidence in ClinVar, or if annotated as P/LP according to ACMG criteria, using the semi-automated CardioClassifier decision support tool.^23^ Other variants that were consistent with known disease mechanisms and sufficiently rare, but not recorded in ClinVar and without other computationally available data to robustly classify as P/LP, were defined as indeterminate sarcomeric variants (**SARC-IND**).^24,25^ The SARC-IND strata differs from those classically termed variants of unknown significance (VUS) as it likely contains additional variants that would be reported as P/LP if subject to full manual curation, which is not feasible in >5000 individuals. Therefore, conservative computational prioritisation was undertaken. Individuals carrying variants that did not fit into these three categories were removed from analyses, including those with rare variants in genes associated with HCM genocopies, those with intermediate frequency variants (FAF>0.00004 or MAF>0.001), and those with variant classes not robustly established as disease-causing (e.g. truncating variants in *MYH7*).^22^ For details on the variant curation pipelines, please see **Supplementary Appendix** (code available on GitHub - details under data availability). Secondary analyses explored whether variants in the SARC-IND category might be further stratified based on features suggested as important but not yet universally adopted in diagnostic practice.^25^ These analyses can be found in the **Supplementary Appendix – SARC-IND sub-group analysis** section.

### Outcome measures

The effect of genotype strata on clinical outcomes was assessed using lifetime risk. The UKBB reports the date of first occurrence of a diagnosis, identified from self-reporting, primary care, hospital in-patient, and death register records. This permitted the identification of events preceding recruitment to the UKBB. The primary clinical outcome was a composite of all-cause mortality or major adverse cardiovascular events (MACE) defined as a diagnostic code for heart failure (including cardiomyopathy), arrhythmia, stroke or cardiac arrest events. Secondary clinical outcomes were the individual components of the primary clinical outcome. Full list of endpoint definitions and data fields used from the UKBB database can be found in the **Supplementary Appendix** and **Table S2**.

### Statistical Analysis

Statistical analysis was performed with R (version 3.6.0) and RStudio Server (version 1.043; Boston, MA), unless otherwise stated. Variables were expressed as percentages if categorical, mean ± standard deviation (SD) if continuous and normal, and median ± inter-quartile range (IQR) if continuous and non-normal. Baseline anthropometric data were compared by Kruskal-Wallis tests and, if differences were identified, a Wilcoxon test was used for pairwise comparisons with Benjamini-Hochberg adjustment for multiple testing. Imaging parameters in two or more groups were compared using analysis of covariance, adjusted for relevant clinical covariates. When differences were significant, a Tukey post-hoc test was applied for pairwise multiple comparisons. Three-dimensional phenotypic regression modelling applied threshold-free cluster enhancement and permutation testing to derive the p values associated with each regression coefficient following adjustment to control the false discovery rate, as previously described.^14,18^

Clinical outcomes were analysed in participants with WES stratified by genotype categories. Cox proportional hazards were calculated for lifetime risk of clinical events, adjusting for sex. For the primary outcome, competing risk analysis was performed using the cause-specific survival method.^26^ Secondary analysis of incident clinical events from recruitment excluded individuals with events preceding enrolment, and was performed using Cox proportional hazards adjusted for age at recruitment and sex. Time-to-event was censored at first event for each outcome, death, or last recorded follow-up. The effect of WT on clinical outcomes was assessed in participants who had undergone CMR. To assess whether genotype-associated risk of clinical events was mediated through structural phenotypes, additional analyses were performed in individuals with WES and CMR data adjusting for maximum WT both as a continuous (per mm increase) and categorical (<13mm, 13-15mm and ≥15mm groups) variable, and left atrial volume, using Cox proportional hazards models. To identify sex-related differences, sex-stratified outcome analysis was carried out. Two sensitivity analyses were performed, excluding patients with known cardiomyopathy from the analysis to ensure observed associations were not driven predominantly by individuals with known disease, and excluding diagnostic codes for cardiomyopathy from the MACE heart failure endpoint, to ensure that associations were with clinical events rather than diagnostic labels. Schoenfeld residuals were checked to confirm assumptions of proportional hazards. Outcomes are reported as hazard ratios (HR) with 95% confidence intervals (CI) and presented graphically as cumulative hazards and Cox proportional hazards curves.

## Results

### Participants

We analysed WES from 200,584 participants and CMR from 39,551 subjects - of which 21,322 had both CMR and WES data available (flowchart in **Figure 2; Table 1**).

**Table 1.** Subject Characteristics and CMR-Derived Cardiac Measurements by genotype: genotype negative (SARC-NEG), carriers of pathogenic or likely pathogenic sarcomeric variants (SARC-P/LP), or carriers of other rare indeterminate sarcomeric variants (SARC-IND). Values are n (%) or mean ± standard deviation. Bold values are statistically significant. Kruskal-Wallis tests were carried out for each variable to determine whether differences in patients’ characteristics between genotype group were significant. If so, a Wilcoxon’s test was used to carry out pairwise comparisons between groups, with Benjamini-Hochberg adjustment for multiple testing, and those p values are shown. For the CMR-derived cardiac parameters analysis was adjusted for age, sex, race, systolic blood pressure (SBP) and body surface area (BSA), using an analysis of covariance (ANCOVA). When differences between genotype groups were significant, a Tukey post-hoc test was applied for pairwise multiple comparisons and those p values are shown. Concentricity = (left ventricular mass / left ventricular end-diastolic volume); CMR = cardiac magnetic resonance; DBP = diastolic blood pressure; EDV = end-diastolic volume; EF = ejection fraction; ESV = end-systolic volume; FD = fractal dimension; HCM = hypertrophic cardiomyopathy; LA = left atrial; LV = left ventricular; LVM = left ventricular mass; peak diastolic strain rate = PDSR; RA = right atrial; RV = right ventricular; SBP = systolic blood pressure; WT = wall thickness.

| Characteristic | SARC-NEG<br>(n =157,922) | SARC-IND<br>(n = 5,253) | SARC-P/LP<br>(n = 474) | SARC-NEG<br>vs<br>SARC-P/LP | SARC-NEG<br>vs<br>SARC-IND | SARC-IND<br>vs<br>SARC-P/LP |
| --- | --- | --- | --- | --- | --- | --- |
| Sex, female | 86,688<br>(54.9%) | 2,884<br>(54.9%) | 265<br>(55.9%) |  |  |  |
| Age, yrs | 56.5 ± 8.1 | 56.3 ± 8.2 | 56.6 ± 8.1 |  |  |  |
| Race, Caucasian | 150,440<br>(85.0 %) | 4,785<br>(79.3%) | 388<br>(81.9%) | 0.09 | **<0.001** | 0.18 |
| SBP, mmHg | 139.7 ± 19.6 | 139.4 ± 19.7 | 139 ± 19.6 |  |  |  |
| DBP, mmHg | 82.2 ± 10.7 | 82.2 ± 10.5 | 81.6 ± 10.9 |  |  |  |
| BSA, m<sup>2</sup> | 1.9 ± 0.22 | 1.9 ± 0.22 | 1.9 ± 0.22 |  |  |  |
| eGFR, mL/min/1.73 m<sup>2</sup> | 90.7 ± 13.4 | 91.1 ± 13.3 | 90.4 ± 13.2 |  |  |  |
| Hypercholesterolaemia | 29,137<br>(18.5%) | 952<br>(18.1%) | 93<br>(19.6%) |  |  |  |
| Hypertension | 52,356<br>(33.2%) | 1,727<br>(32.9%) | 168<br>(35.4%) |  |  |  |
| Diabetes | 8,429<br>(5.3%) | 313<br>(6.0%) | 23<br>(4.9%) |  |  |  |
| Aortic valve disease | 1,567<br>(1.0%) | 38<br>(0.7%) | 7<br>(1.5%) |  |  |  |
| Known HCM | 109<br>(0.07%) | 7<br>(0.1%) | 20<br>(4.2%) | **<0.001** | 0.09 | **<0.001** |
| On medication for<br>cholesterol, blood<br>pressure, diabetes | 18,537<br>(11.7%) | 606<br>(11.5%) | 59<br>(12.5%) |  |  |  |

**Table 1** - Subject Characteristics and CMR-Derived Cardiac Measurements by genotype: genotype negative (SARC-NEG), carriers of pathogenic or likely pathogenic sarcomeric variants (SARC-P/LP), or carriers of other rare indeterminate sarcomeric variants (SARC-IND).
|  | SARC-NEG<br>(n = 16,883) | SARC-IND<br>(n = 521) | SARC-P/LP<br>(n = 43) | SARC-NEG<br>vs<br>SARC-P/LP | SARC-NEG<br>vs<br>SARC-IND | SARC-IND<br>vs<br>SARC-P/LP |
| --- | --- | --- | --- | --- | --- | --- |
| LVEDV, mL | 147.9 ± 33.5 | 149.9 ± 34.5 | 140.5 ± 31.2 | 0.14 | **0.004** | **0.02** |
| LVESV, mL | 60.4 ± 19 | 61.4 ± 19.7 | 55.5 ± 16.4 | 0.08 | **0.04** | **0.02** |
| LVEF, % | 59.5 ± 6.1 | 59.5 ± 6.1 | 60.8 ± 6.1 |  |  |  |
| LVM, g | 86 ± 22.1 | 87 ± 23.5 | 88.6 ± 27.5 | 0.25 | **0.001** | 0.86 |
| LV Max WT, mm | 9.4 ± 1.6 | 9.5 ± 1.7 | 10.9 ± 2.7 | **<0.001** | **0.002** | **<0.001** |
| LV Concentricity, g/mL | 0.58 ± 0.09 | 0.58 ± 0.09 | 0.63 ± 0.12 | **<0.001** | 0.92 | **<0.001** |
| LV Global longitudinal strain, % | -18.5 ± 2.8 | -18.5 ± 2.8 | -18.9 ± 2.4 |  |  |  |
| LV Global radial strain, % | 45 ± 8.3 | 44.9 ± 8.2 | 45.6 ± 8.5 |  |  |  |
| LV Global circumferential strain, % | -22.3 ± 3.4 | -22.2 ± 3.3 | -22.4 ± 3.2 |  |  |  |
| LV Longitudinal PDSR | 1.7 ± 0.6 | 1.6 ± 0.6 | 1.7 ± 0.6 |  |  |  |
| LV Radial PDSR | -5.7 ± 2 | -5.7 ± 1.9 | -5.9 ± 2 |  |  |  |
| LAEDV, mL | 72.7 ± 23.1 | 73.7 ± 27.8 | 81 ± 27.1 | **0.02** | 0.2182 | 0.08 |
| LAESV, mL | 29.2 ± 15.3 | 30.3 ± 20.7 | 33.6 ± 16 | 0.0984 | 0.1358 | 0.31 |
| LAEF, % | 61.3 ± 9.2 | 60.9 ± 9.8 | 59.8 ± 8.6 |  |  |  |
| RVEDV, mL | 156.6 ± 37.4 | 157.4 ± 37.5 | 139.6 ± 33.4 | **<0.001** | 0.09 | **<0.001** |
| RVESV, mL | 67.6 ± 21.3 | 67.7 ± 21.1 | 56.6 ± 15.4 | **<0.001** | 0.44 | **<0.001** |
| RVEF, % | 57.3 ± 6.2 | 57.4 ± 6 | 59.7 ± 5.1 | **0.007** | 0.9 | **0.02** |
| RAEDV, mL | 86 ± 27.7 | 88.9 ± 32.8 | 81.4 ± 28.7 | 0.38 | **0.009** | 0.09 |
| RAESV, mL | 46.1 ± 19.1 | 47.7 ± 24.3 | 43.1 ± 19.4 | 0.39 | **0.05** | 0.13 |
| RAEF, % | 47.1 ± 9.4 | 47.6 ± 9.9 | 48.2 ± 9.5 |  |  |  |
| Mean apical FD | 1.14 ± 0.04 | 1.14 ± 0.04 | 1.16 ± 0.05 | **0.006** | 0.88 | **0.01** |
| Mean basal FD | 1.19 ± 0.03 | 1.19 ± 0.03 | 1.2 ± 0.04 |  |  |  |
| Mean Global FD | 1.17 ± 0.03 | 1.17 ± 0.03 | 1.19 ± 0.04 | **0.003** | 0.93 | **0.006** |

**Figure 2.**
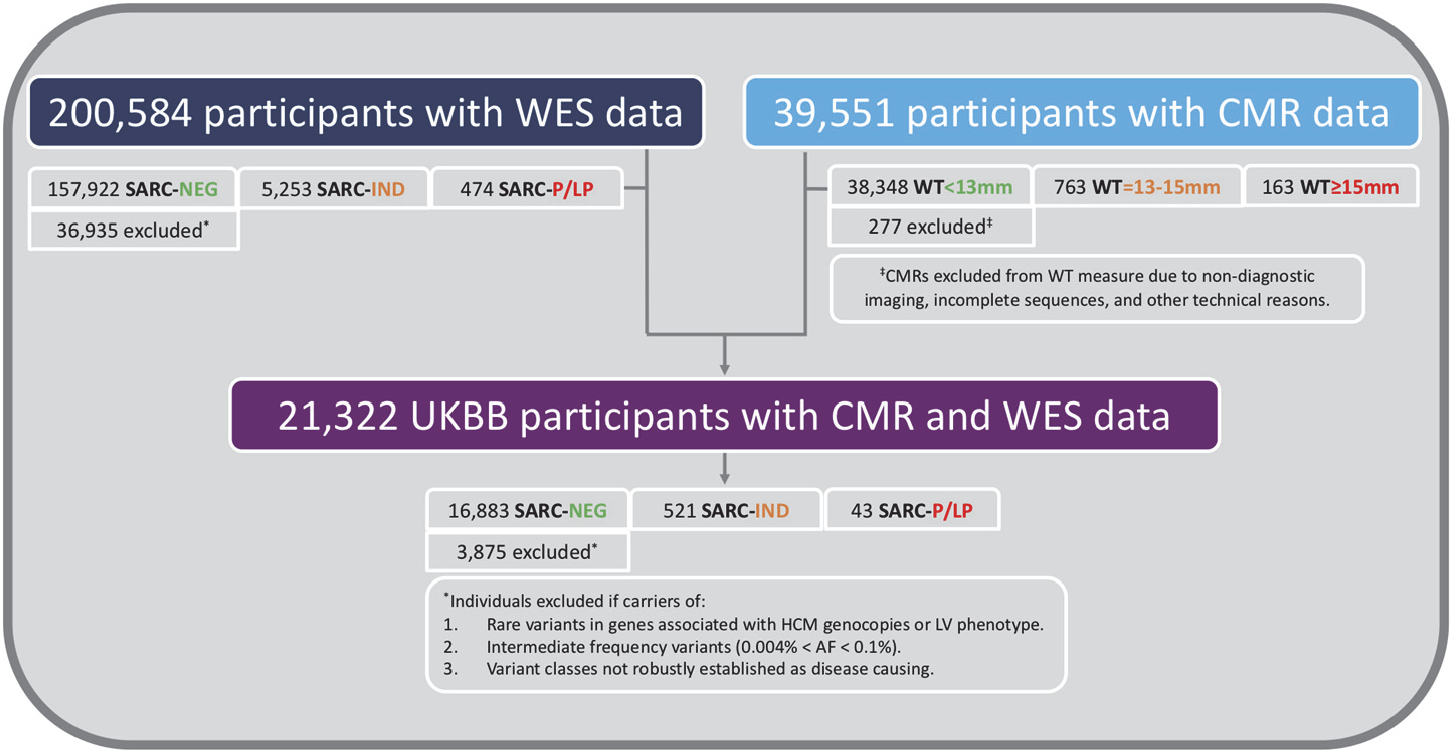
Flow chart of 218,813 UKBB participants included in this study. CMR = cardiac magnetic resonance; HCM = hypertrophic cardiomyopathy; AF = allele frequency; SARC-NEG = genotype negative; SARC-IND = carriers of indeterminate sarcomeric variants; SARC-P/LP = carriers of pathogenic or likely pathogenic sarcomeric variants; UKBB = UK Biobank; WES = whole exome sequencing; WT = wall thickness

### Prevalence of rare sarcomeric variants

There were 157,922 SARC-NEG subjects, 474 (0.24%) carriers of a heterozygous SARC-P/LP variant, and 5,253 (2.6%) carriers of SARC-IND variants, including 113 compound heterozygotes where neither variant met criteria to be classified SARC-P/LP (**Table S3)**.

### Left ventricular wall thickness in the general population

Out of 39,274 subjects that underwent CMR analysis, 763 (1.9%, 1 in 51) had mild hypertrophy (WT 13-15mm) and 163 (0.4%, 1 in 241) had at least moderate hypertrophy (WT ≥15mm). Participants with WT ≥13mm were older (66.3±7.4 vs 63.6±7.6 years, p<0.001), more often males (88.4% vs 46.9% male, p<0.001), with higher systolic blood pressure (SBP) (149.5.2±18.8 vs 138±18.2 mmHg, p<0.001) and body surface area (BSA) (2.07±0.21 vs 1.86±0.21 m^2^, p<0.001). There were no differences in terms of race. The prevalence of phenotypic HCM, defined as WT ≥15mm in the absence of hypertension and valve disease, was 0.19% (n=76, 1 in 517).

### Penetrance and expressivity of rare sarcomeric variants

Analyses were restricted to UKBB participants with both CMR and WES (n=21,322). In this subset 16,883 were denoted as SARC-NEG, 521 as SARC-IND and 43 as SARC-P/LP. CMR-derived cardiac measurements by genotype are summarised in **Table 1**. Wall thickness was greater in SARC-P/LP versus SARC-NEG (**Figure 3**) and marginally greater in SARC-IND after adjustment for relevant clinical variables (**Table 1**). When compared to SARC-NEG, SARC-P/LP show evidence of concentric remodelling (increased mass / volume ratio), smaller right ventricular volumes, higher left atrial volume and increased trabeculation (**Table 1)**. Independent, 3D analysis of cardiac geometry (**Figure 3**) showed that SARC-IND were positively associated with increased WT (n=516, mean β=0.08, significant area=71.2%), predominantly across the mid-basal anterior septum in continuity with the anterior wall. There was a much stronger positive association between SARC-P/LP and increased WT (n=42, mean β=0.32, significant area=50.2%) involving most of the septum, anterior wall and apex.

**Figure 3.**
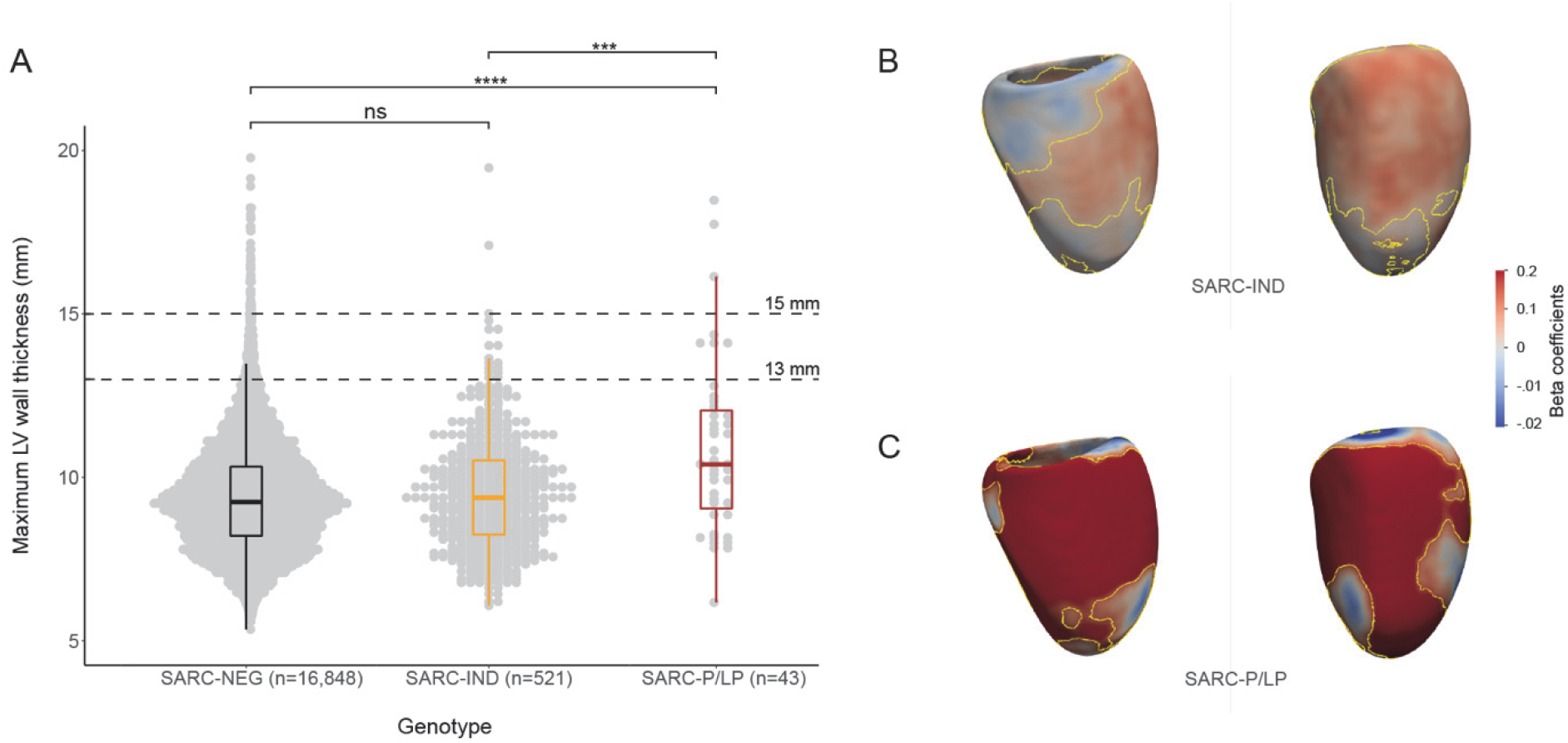
Dot and boxplots of maximum wall thickness by genotype (A): genotype negative (SARC-NEG), carriers of indeterminate sarcomeric variants (SARC-IND) or of pathogenic or likely pathogenic sarcomeric variants (SARC-P/LP). Also shown are the wall thickness thresholds of 15mm, which defines HCM if no other cause of hypertrophy is found, and 13mm, above which the diagnosis of HCM can be made in first-degree relatives of someone with unequivocal HCM. ** = p<0.01; *** = p <= 0.001; **** = p <= 0.0001. Three-dimensional parametric modelling of left ventricular geometry in 17,219 UK Bioban participants with whole exome sequencing (B and C). The models represent the surface of the left ventricle on which standardised beta coefficients are mapped to show the regional strength of association between genotype status and wall thickness with adjustment for age, gender, systolic blood pressure, body surface area and race. SARC-IND (n=516) were weakly associated with increased wall thickness in the septum (mean β = 0.08, significant area = 71.2%), while SARC-P/LP had a much stronger positive association (n=42; mean β = 0.32, significant area = 50.2%). Contour lines indicate significant regions (p<0.05) after correction for multiple testing. LV projections are en-face septal (left) and anterior (right) walls.

In the 43 SARC-P/LP variant carriers, LV hypertrophy (≥13mm) was present in 7, corresponding to a penetrance of 16% (CI 7-31%) for this phenotype. Of the 521 carriers of SARC-IND variants, 17 individuals (3%, CI 1.9-5%) had WT ≥13mm. In 391 out of these 564 individuals with a SARC-IND or SARC-P/LP variant, there was no concomitant hypertension or valve disease (69.3%). Amongst these 391 individuals, the penetrance of LVH ≥13mm was 6.5% for SARC-P/LP (2/31), and 1% for SARC-IND (4/360). Details of variants found in these six individuals with otherwise unexplained LVH are shown in **Table S4**. Amongst the 173 individuals with a rare sarcomere-encoding variant and hypertension or valve disease, the penetrance of LVH was 42% for SARC-P/LP (5/12), and 8% for SARC-IND (13/161). Only 1 individual out of the 40 with a wall thickness ≥15mm (2.5%), not accounted for by hypertension or valve disease, carried a SARC-P/LP variant.

### Clinical outcomes associated with rare sarcomeric variants

Clinical outcomes for 163,649 participants were analysed. Median age at recruitment was 58 (IQR 50-63 years), and participants were followed up for a median of 10.8 years (IQR 9.9-11.6 years) with total of 19,507 primary clinical events reported. Among SARC-NEG (n=157,922), SARC-IND (n=5,253) and SARC-P/LP (n=474) individuals, there were 18,793 (11.9%), 621 (11.8%) and 93 (19.6%) primary clinical outcome events (all-cause mortality or MACE), respectively.

Examining lifetime risks, SARC-P/LP variants were associated with increased risk of death or MACE (HR 1.69, 95% CI 1.36-2.09, p<0.001), heart failure (HR 4.40, 95% CI 3.22-6.02, p<0.001) and arrhythmia (HR 1.55, 95% CI 1.18-2.03, p=0.002) (**Figure 4, Table S5**). There was no difference in risk in any of the primary or secondary clinical outcomes when comparing SARC-IND and SARC-NEG. Sex was independently associated with all clinical outcomes, with men having an increased risk. While males have a higher overall risk of adverse outcomes (HR 1.76, CI 1.71-1.81 p<0.001), the incremental genetic risk from SARC-P/LP mutations compared with SARC-NEG is greater in females (HR for females: 2.18 CI 1.65-2.89, p<0.001; HR for males: 1.42 CI 1.05-1.9, p=0.02; HR for sex*SARC-P/LP interaction: 1.55, CI 1.03-2.33, p=0.04) (**Table S6; Figure S1**).

**Figure 4.**
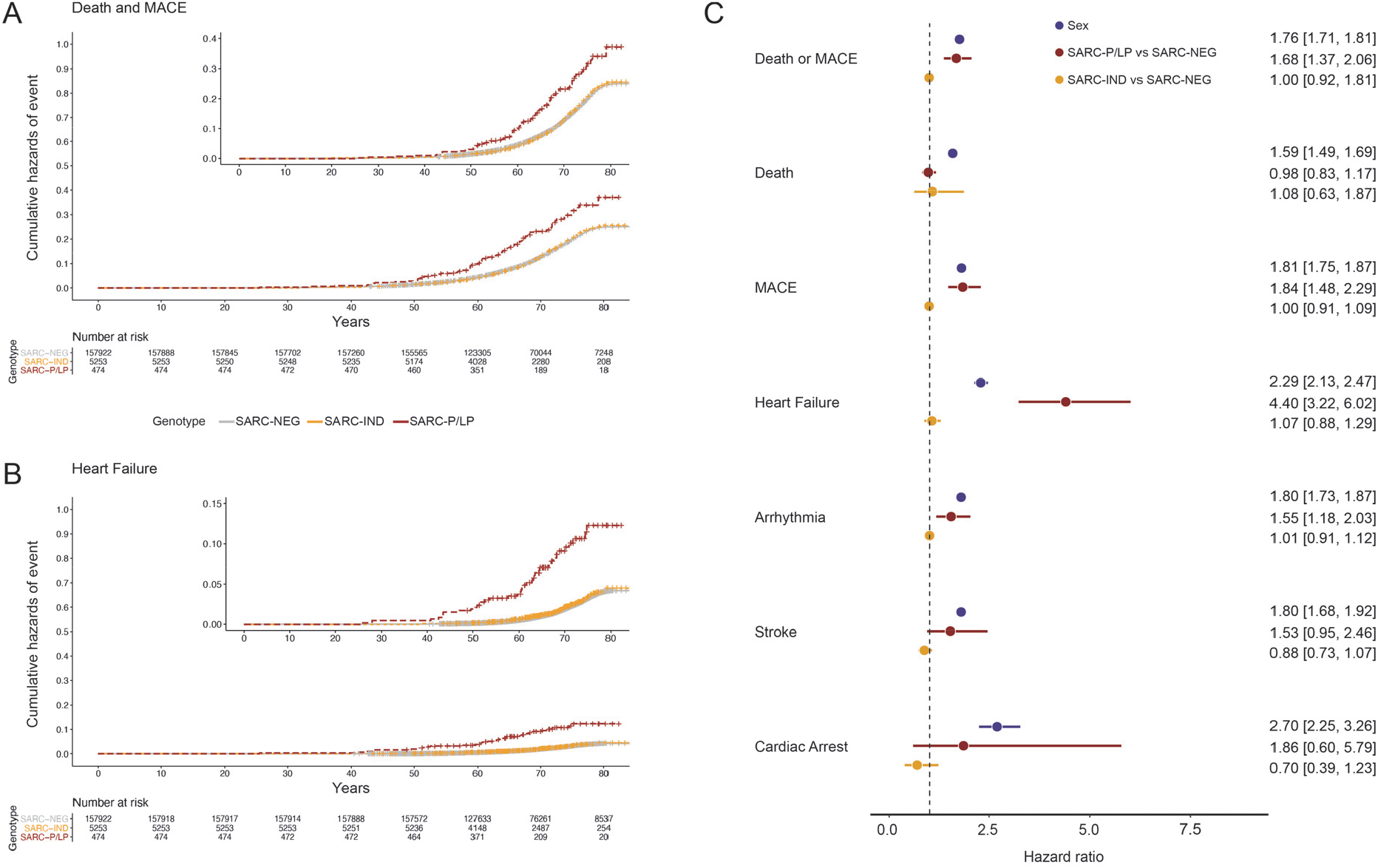
Cumulative hazard curves with zoomed plots for lifetime risk of A) death and major adverse cardiovascular events (MACE; consisting of heart failure, arrhythmia, stroke and cardiac arrest events) or B) heart failure, stratified by genotype: genotype negative (SARC-NEG), carriers of indeterminate sarcomeric variants (SARC-IND) or of pathogenic or likely pathogenic sarcomeric variants (SARC-P/LP). C) Forest plot of comparative lifetime risk of clinical endpoints (Cox proportional hazards models adjusted for sex) by genotype. Sex refers to males compared to females.

Examining incident risks, primary clinical events were increased in SARC-P/LP (HR 1.54, CI 1.18-2.01, p=0.001) but not in SARC-IND (**Table S7**). Sensitivity analyses, excluding patients with cardiomyopathy, and excluding cardiomyopathy as an outcome, yielded similar results to the primary analysis (**Table S8**) confirming that previous findings were neither driven by individuals with known disease, nor by a diagnostic label of cardiomyopathy as an endpoint.

In the 39,551 individuals that underwent CMR, the number of primary clinical events was 2,568 (6.7%), 112 (14.7%) and 34 (20.9%) in the WT <13mm, 13-15mm and ≥15mm sub-groups, respectively. Increased WT was associated with increased risk of death or MACE (per mm increase: HR 1.15, 95% CI 1.11-1.20, p<0.001) and each individual endpoint (**Table S9; Figure S2**), with similar findings in the imaging and WES sub-group (n=17,447) (**Table S10; Figure S3**). Increased trabecular complexity, as measured by FD analysis, was associated with adverse clinical outcomes (HR per 0.1 unit increase in mean global FD 1.31, 95% CI 1.12-1.54, p=0.001) after adjustment for SBP and WT (**Table S11; Figure S4**).

Among 17,447 individuals with imaging and WES data, there were 1,178 (7.0%), 45 (8.4%) and 7 (15.9%) primary clinical events in SARC-NEG, SARC-IND and SARC-P/LP, respectively. After adjustment for WT, the effect of genotype on clinical outcomes was reduced but persisted (**Table S12**). The risk of heart failure remained higher in SARC-P/LP (HR 5.25, 95% CI 1.62-17, p=0.02) and SARC-IND (HR 2.21, CI 1.16-4.23, p=0.006) despite adjustment for WT and left atrial volume (**Table S13**).

## Discussion

In a study of over 200,000 adults, we found the prevalence of rare variants in HCM-associated sarcomere-encoding genes to be 2.9% with the prevalence of SARC-P/LP variants conservatively measured at 0.24%. The previous largest investigation,^27^ gave the prevalence of pathogenic / likely pathogenic HCM variants in 3,600 participants of the Framingham Heart Study and Jackson Heart Study as 0.6%, with 11.2% of individuals having one or more rare variant in a sarcomere-encoding gene, but at a less conservative frequency threshold and prior to contemporary variant classification guidelines.^24^

We found that SARC-P/LP variants were associated with an increased lifetime risk of adverse cardiovascular outcomes, predominantly due to heart failure and arrhythmia, that was independent of LV wall thickness – the cardinal diagnostic feature of HCM. Such variants are also associated with greater risk in HCM patient cohorts, with adverse outcomes reported to be twice as likely in HCM patients carrying SARC-P/LP variants.^2^ SARC-P/LP variants, were associated with a greater increase in lifetime risk of death and MACE in females than in males. This supports findings in HCM patients, where despite apparent reduced disease penetrance,^28^women have lower survival, regardless of genotype.^29,30^ In contrast, SARC-IND variant carriers, which include a mixture of benign variants and variants with pathogenic potential, express a minimal phenotype and have a generally benign clinical course.

Adults carrying SARC-P/LP variants had a sub-clinical phenotype characterised by increased anteroseptal wall thickness, concentric ventricular remodelling, increased left atrial volume and greater trabeculation. In SARC-P/LP variant carriers, hypertrophy (≥13mm) was present in 16% of individuals. Penetrance estimates from family studies vary, but a recent study of relatives of HCM probands observed an HCM phenotype in 37% of genotype positive relatives at first screening (mean age 41), with ∼30% of the remainder manifesting a phenotype by age 60.^31^ Although UKBB participants have a higher degree of kinship than would be expected in a random sample,^32^ these results suggest that the penetrance of familial HCM is additionally driven by other genetic or environmental disease modifying characteristics, shared within families but which overlap less commonly in the community. Conversely, only 2.5% of individuals with unexplained LVH (WT≥15mm) were carriers of SARC-P/LP variants. This contrasts with the diagnostic yield in HCM cohorts, where even patients without family history have a comparatively high 30% yield of sarcomere variants,^33^suggesting that patients enrolled into cohorts and/or referred for diagnostic sequencing represent a skewed subset, likely enriched by those with clinical risk factors.^34,35^These findings emphasise the importance of combined clinical and genetic assessment in risk stratifying patients with unexplained LV hypertrophy and managing the appropriate screening of relatives.^6^

We determined the prevalence of unexplained hypertrophy (WT≥15mm in the absence of hypertension and valve disease) as 1 in 517, confirming historical estimates of HCM prevalence observed in young adults using echocardiography.^3^However, this is a conservative estimate as hypertension and valve disease, of unknown degree of severity, were present in 34% of all participants and might not be sufficient to explain the degree of LV hypertrophy. If not accounting for this, at least moderate hypertrophy (WT≥15mm) was detected in 1 in 241 individuals. Increasing WT was associated with higher risk of death and MACE (and each individual component), which persisted despite adjustment for sex, sarcomeric genotype and systolic blood pressure.

In conclusion, we found that SARC-P/LP variants are present in 1 in 423 adults. Although the presence of SARC-P/LP variants in individuals in the community was rarely associated with the degree of unexplained hypertrophy required for a diagnosis of HCM, they are associated with a sub-clinical cardiomyopathic phenotype and an increased risk of adverse cardiovascular outcomes. The presence of SARC-P/LP variants may be of clinical relevance to >18 million people worldwide and, with increasing availability of genetic sequencing and the emergence of disease modifying therapies,^36^there is the potential for sarcomeric rare variant analysis to improve cardiovascular risk assessment and management even when disease is not manifest.

## Data Availability

All data in this study is available from UK Biobank (http://www.ukbiobank.ac.uk/).
Algorithms for imaging and statistical analysis are publicly available at https://github.com/ImperialCollegeLondon/HCM_expressivity (doi:10.5281/zenodo.4428740). 

https://github.com/ImperialCollegeLondon/HCM_expressivity

## Abbreviations

ACMG: American College of Medical Genetics and Genomics
AF: atrial fibrillation
CC: CardioClassifier
CMR: cardiovascular magnetic resonance imaging
DBP: diastolic blood pressure
FAF: filtering allele frequency
FD: fractal dimension
HCM: hypertrophic cardiomyopathy
LV: left ventricle
LVEDV: left ventricular end-diastolic volume
LVEF: left ventricular ejection fraction
LVESV: left ventricular end-systolic volume
LVH: left-ventricular hypertrophy
MAF: minor allele frequency
SARC-P/LP: pathogenic or likely pathogenic variants in sarcomere-encoding genes
SARC-NEG: genotype negative / no rare variants in sarcomere-encoding genes
SARC-IND: indeterminate variants in HCM-associated sarcomere-encoding genes (rare variants that do not meet criteria for pathogenic/likely pathogenic annotation)
SBP: systolic blood pressure
UKBB: UK Biobank
WT: wall thickness

## Data Access

All data in this study is available from UK Biobank (http://www.ukbiobank.ac.uk/).

Algorithms for imaging and statistical analysis are publicly available at https://github.com/ImperialCollegeLondon/HCM_expressivity (doi:10.5281/zenodo.4428740).

