## Supplementary Appendix for "Outcomes and phenotypic expression of rare variants in hypertrophic cardiomyopathy genes amongst UK Biobank participants"

### Supplementary Table 1: STROBE Statement

#### Checklist of items that should be included in reports of ***cohort studies***

| | Item No | Recommendation | Page number |
|---|---|---|---|
| **Title and abstract** | 1 | (*a*) Indicate the study's design with a commonly used term in the title or the abstract | p.1 |
| | | (*b*) Provide in the abstract an informative and balanced summary of what was done and what was found | p.2 |
| **Introduction** | | | |
| Background/rationale | 2 | Explain the scientific background and rationale for the investigation being reported | p.4 |
| Objectives | 3 | State specific objectives, including any prespecified hypotheses | p.4 |
| **Methods** | | | |
| Study design | 4 | Present key elements of study design early in the paper | p.4-5 |
| Setting | 5 | Describe the setting, locations, and relevant dates, including periods of recruitment, exposure, follow-up, and data collection | p.5 |
| Participants | 6 | (*a*) Give the eligibility criteria, and the sources and methods of selection of participants. Describe methods of follow-up | p.5-6 and Fig 2 |
| | | (*b*) For matched studies, give matching criteria and number of exposed and unexposed | NA |
| Variables | 7 | Clearly define all outcomes, exposures, predictors, potential confounders, and effect modifiers. Give diagnostic criteria, if applicable | p.5-8 |
| Data sources/ measurement | 8* | For each variable of interest, give sources of data and details of methods of assessment (measurement). Describe comparability of assessment methods if there is more than one group | p.5-8 and sup appendix as indicated for each in the main manuscript |
| Bias | 9 | Describe any efforts to address potential sources of bias | p.5-8 |
| Study size | 10 | Explain how the study size was arrived at | p.4-5 |
| Quantitative variables | 11 | Explain how quantitative variables were handled in the analyses. If applicable, describe which groupings were chosen and why | p.7-9 |
| Statistical methods | 12 | (*a*) Describe all statistical methods, including those used to control for confounding | p.8-9 |
| | | (*b*) Describe any methods used to examine subgroups and interactions | p.8-9 and then and sup appendix as indicated for each in the main manuscript |
| | | (*c*) Explain how missing data were addressed | NA |
| | | (*d*) If applicable, explain how loss to follow-up was addressed | NA |
| | | (*e*) Describe any sensitivity analyses | p.8 and Table S8 |
| **Results** | | | |

| | | | |
|---|---|---|---|
| Participants | 13* | (a) Report numbers of individuals at each stage of study—eg numbers potentially eligible, examined for eligibility, confirmed eligible, included in the study, completing follow-up, and analysed | p.8 in particular, then p.8-11 including reference to Table 1 and Figure 2 |
| | | (b) Give reasons for non-participation at each stage | p.6-7 and Figure 2 |
| | | (c) Consider use of a flow diagram | Figure 2 |
| Descriptive data | 14* | (a) Give characteristics of study participants (eg demographic, clinical, social) and information on exposures and potential confounders | p.8 referring to Table 1 |
| | | (b) Indicate number of participants with missing data for each variable of interest | p.8 and Figure 2 |
| | | (c) Summarise follow-up time (eg, average and total amount) | p.10 |
| Outcome data | 15* | Report numbers of outcome events or summary measures over time | p.10-11 |
| Main results | 16 | (*a*) Give unadjusted estimates and, if applicable, confounder-adjusted estimates and their precision (eg, 95% confidence interval). Make clear which confounders were adjusted for and why they were included | p.8-11, table 1, figure 3 and 4 |
| | | (*b*) Report category boundaries when continuous variables were categorized | p.8-9 |
| | | (*c*) If relevant, consider translating estimates of relative risk into absolute risk for a meaningful time period | NA |
| Other analyses | 17 | Report other analyses done—eg analyses of subgroups and interactions, and sensitivity analyses | p.7 referring to sub-group analysis section in the sup appendix; p.10-11 |
| **Discussion** | | | |
| Key results | 18 | Summarise key results with reference to study objectives | p.11-14 |
| Limitations | 19 | Discuss limitations of the study, taking into account sources of potential bias or imprecision. Discuss both direction and magnitude of any potential bias | p.13 |
| Interpretation | 20 | Give a cautious overall interpretation of results considering objectives, limitations, multiplicity of analyses, results from similar studies, and other relevant evidence | p.11-13 |
| Generalisability | 21 | Discuss the generalisability (external validity) of the study results | p.13-14 |
| **Other information** | | | |
| Funding | 22 | Give the source of funding and the role of the funders for the present study and, if applicable, for the original study on which the present article is based | p.14 |

*Give information separately for exposed and unexposed groups.

#### Three-dimensional phenotyping

Three-dimensional parametric analysis of cardiac geometry to model the expressivity of genetic variants has been described previously.[1,2] Briefly, we performed a high resolution segmentation of the heart using a deep learning neural network algorithm that was trained on over 2000 three-dimensional CMRs, enabling spatial consistency without compromising segmentation accuracy.[3] This method is robust and capable of producing accurate, high-resolution, and anatomically smooth bi-ventricular models, despite the presence of artifacts on the original CMR datasets.[4] The models are then registered to a standard reference template (cardiac atlas) so that precise comparisons can be made within and between populations. A general linear model is fitted at each vertex of the left ventricular mesh with wall thickness as the dependent variable and genotype status as the independent variable with demographic and physiological parameters as co-variates. Threshold-free cluster enhancement (TFCE) is then applied to boost belief in extended areas of coherent signal. The permutation testing procedure employed by this approach is the Freedman-Lane procedure, whilst a false discovery rate (FDR) correction using the Benjamini-Hochberg procedure is applied to correct for multiple testing.

#### Cardiac motion analysis

Motion tracking was performed using non-rigid image registration and to reduce the accumulation of registration errors this was performed in both forward and backward directions from the end-diastolic frame and an average displacement field calculated. This motion field was then used to warp the segmentation contours from end-diastole onto successive adjacent frames. Circumferential ($E_{cc}$) and radial ($E_{rr}$) strains were calculated on the short axis cines by the change in length of respective line segments over time. Motion tracking was also performed on the long-axis four-chamber cines to derive longitudinal ($E_{ll}$) strain. Peak strain for each segment and global peak strain were then calculated. Strain was measured from slices acquired at basal, mid-ventricular, and apical levels. Strain rate was estimated as the first derivative of strain and peak early diastolic strain rate in radial ($PDSR_{rr}$) and longitudinal ($PDSR_{ll}$) directions was detected using an algorithm to identify local maxima.

#### Variant curation pipeline

This pipeline can be found on GitHub (See **Data Access** in main manuscript). The UKBB exome data was annotated using Ensembl Variant Effect Predictor (VEP; version 99.1)[5] with plugins for ClinVar (version 20201026)[6], gnomAD (version r2.1)[7], dbscSNV[8], MaxEntScan[9] and LOFTEE.[7] The VEP output was analysed using R (version 3.6.0) and Rstudio (version 1.3.1073).

Canonical, protein altering variants that had a MAF of <0.1% in gnomAD and UKBB were included in the analyses. Protein altering variants were specified as high or moderate impact by Sequence Ontology and ENSEMBL, with the addition of splice region variants for further curation. The variants were filtered for genes and protein consequences of interest (as described by Ingles et al. [10]) to include 8 definitive-evidence sarcomeric HCM genes (*MYH7, MYBPC3, MYL2, MYL3, ACTC1, TNNI3, TNNT2, TPM1*), 3 medium-evidence HCM genes (*CSRP3, TNNC1, JPH2*), 2 intrinsic cardiomyopathy genes (*ACTN2* (moderate classification), *PLN* (definitive classification)), and 12 syndromic genes that can cause isolated left ventricular hypertrophy (*FHL1, TTR, FLNC, GLA, LAMP2, PRKAG2, PTPN11, RAF1, RIT1, ALPK3, CACNA1C, DES*).

Splice region variants (outside the canonical splice donor & acceptor sites) were assessed in two ways; i) via ClinVar report: splice region variants found pathogenic with at least 2 star evidence for HCM in ClinVar and reported functional evidence for splicing were termed “splice confirmed”; if the functional evidence was unclear the protein consequence remained unchanged; if there was functional evidence of an alternative mechanism to splicing, the protein consequence was renamed (e.g. missense variant); ii) via prediction threshold: of the HCM-pathogenic splice confirmed variants, all were found to meet the following MaxEnt and dbSNV thresholds; MaxEntScan difference > 3, ADA score > 0.95, and RF score > 0.75, and these thresholds were used to identify potentially splice-causing variants of those splice region variants identified with a non-synonymous consequence flag (e.g. intron variant).

The pipeline then consisted of four main filtering steps which resulted in an output of four columns of binary code flagging carriers (heterozygous, compound heterozygotes, and homozygotes, combined) as “1”:

SARC-NEG – Identification of carriers of any rare non-synonymous variants in any of the 25 genes of interest: too inclusive to expect a significant signal but identified genotype negatives.

SARC-IND – Identification of carriers of gene-specific consequences of the 8 sarcomeric genes: this step separated the variants into two subsets; i) Loss of function (LoF) alleles (deemed group A), which contained only the gene *MYBPC3*, and filters for the protein consequences of stop_gained, splice_acceptor_variant, splice_donor_variant, frameshift_variant, and splice_region_variant (with additional in silico evidence of an effect on splicing). LOFTEE was incorporated in this step to exclude LoF variants that were flagged as “low confidence” (LC) and other LOFTEE flags, such as “NAGNAG site” requiring reannotation to non-LoF variant status; ii) Protein altering (PAV) alleles (deemed group B), which included all 8 sarcomeric genes, including *MYBPC3*, and filters for the protein consequences of missense_variant, inframe_insertion, and inframe_deletion. Both groups included additional positional annotation (LoF variants found in the last exon or 55bp into the penultimate exon), this included variants that introduce a protein-truncating variant (PTC) and predicted to lead to nonsense-mediated decay (NMD). The variants flagged “coding sequence variant” and “protein altering variant” were manually curated, as were “stop_lost” and “start_lost” which were examined via ENSEMBL sequence and UCSC Genome Browser to identify in-frame rescues nearby. To be included in the SARC-IND group, the variants were required to meet a maximum gnomAD filter allele frequency (FAF) threshold for HCM (<0.00004).[11] The SARC-IND variant group is like that of the standard flag “VUS” for variants of unknown significance, except that it likely contains additional variants that would be reported as pathogenic/likelypathogenic if subject to full manual curation, which is not feasible with >5,000 individuals; hence a conservative computational prioritisation was undertaken.

SARC-P/LP is as SARC-IND, plus annotated as pathogenic/likely pathogenic according to the ACMG guidelines[12] via CardioClassifier software[11] with additional segregation information (Table S6 in Walsh)[13] or has been reported by multiple submitters, an expert panel, or as practice guidelines (2+ star evidence), in patients with HCM and deemed pathogenic/likely pathogenic based on the evidence provided to ClinVar.

#### Rare variant analysis

In total 157,922 UKBB participants were classified as SARC-NEG and 5,727 participants (129 compound heterozygotes) were carriers of 1,569 variants identified in the 8 sarcomeric genes with predicted protein consequences (e.g. missense variants in all 8 genes plus loss of function variants in *MYBPC3*) and a gnomAD filtering allele frequency for HCM (<0.00004). Of these 1,569 variants, 82 were identified as SARC-P/LP following ACMG guidelines via CardioClassifier software[14] with additional segregation information (Table S6 in Walsh et al)[13], being identified in 404 heterozygous carriers. 67 of the 1,569 variants, were identified as SARC-P/LP with 2-star evidence (multiple submitters) reported on ClinVar and were found in 326 heterozygous carriers. In summary, 474 participants (0.24%, 1 in 423) were heterozygous carriers of a SARC-P/LP variant, and 5,253 participants (2.6%, 1 in 38) were carriers (including 113 compound heterozygotes) of other rare sarcomeric variants of indeterminate pathogenicity (SARC-IND).

#### Outcome analysis - additional methods (R packages)

Statistical analysis was performed using the *survival* and *cmprsk* packages in R. Outcomes are reported as hazard ratios (HR) with 95% confidence intervals (CI) and presented graphically as cumulative hazards and Cox proportional hazards curves using *survminer* package in R.

#### UK Biobank clinical event definitions, data fields and data Sources

##### First occurrences data in the UKBB

All individuals in the UKBB have their health records linked. Data sources for clinical events and diagnoses include hospital in-patient records, death records, cancer register and primary care records. Participants also self-reported diagnoses and estimated diagnoses dates at study recruitment, although this data has not been clinically verified or validated. Diagnostic data from the various sources are mapped to ICD codes, and the date of first diagnosis made available as 'first occurrence' data (Category 1712) (https://biobank.ndph.ox.ac.uk/showcase/showcase/docs/first_occurrences_outcomes.pdf).

Incorporation of diagnostic data across sources that use different data coding methods occurs using mapping of read codes on to a core spine of ICD-10 diagnostic codes using the NHS Digital Technology Reference Data Update Distribution (TRUD: https://isd.digital.nhs.uk/trud3/user/guest/group/0/pack/9) and SNOMED CT (NHS Digital SNOMED-CT: https://digital.nhs.uk/snomed-ct). ICD-10 codes were then selected in first occurrences data.

##### Clinical event definitions

The primary clinical outcome was a composite of major adverse cardiovascular events (MACE) and any cause death. MACE was defined as:

- Heart failure events: heart failure (ICD code: I50, Field ID: 131354), cardiomyopathy (I42, 131338) and cardiomyopathy in diseases classified elsewhere (I43, 131340)
- Arrhythmia events: atrial fibrillation and flutter (I48, 131350), other cardiac arrhythmias (I49, 131352), atrioventricular and left bundle-branch block (I44, 131342), other conduction disorders (I45, 131345), paroxysmal tachycardia (I47, 131348)

- Stroke events: cerebral infarction (I63, 131366)
- Cardiac arrest (I46, 131346)

Secondary clinical outcomes included the individual components of the primary clinical outcome.

#### Supplementary Table 2 – UKBB Fields used in the analyses

| ICD10 | ICD9 | Self-Reported | Operation Code | Operative Procedure Code | Condition |
|---|---|---|---|---|---|
| E780 | 27202 | | | | pure hypercholesterolaemia |
| E782 | | | | | mixed hypercholesterolaemia |
| | 27209 | | | | pure hypercholesterolaemia (other) |
| | 27200 | | | | familial hypercholesterolaemia |
| | 2720 | | | | hypercholesterolaemia |
| | | 1473 | | | high cholesterol |
| | | 1220 | | | diabetes |
| | | 1223 | | | type-2 diabetes |
| | | 1222 | | | type-1 diabetes |
| | | 1490 | | | aortic stenosis |
| | | 1586 | | | aortic valve disease |
| I350 | | | | | Aortic (valve) stenosis |
| I359 | 4241 | | | | Aortic valve disorder, unspecified |
| | | | 1099 | | aortic valve repair/replacement |
| | | | 1097 | | heart valve surgery |
| | | | 1101 | | other valve repair/replacement |
| Q23 | | | | | Congenital malformations of aortic and mitral valves |
| I391 | | | | | Aortic valve disorders in diseases classified elsewhere |
| I060 | 3950 | | | | Rheumatic aortic stenosis |
| Q230 | 7463 | | | | Congenital stenosis of aortic valve |
| I358 | | | | | Other aortic Valve disorders |
| I35 | | | | | Non-rheumatic aortic valve disorders |
| I062 | 3952 | | | | Rheumatic aortic stenosis with insufficiency |
| | 74722 | | | | Other anomalies of aorta (supra-aortic stenosis) |
| | | | | K374 | repair of supra-aortic stenosis |
| Q231 | 7464 | | | | Congenital insufficiency of aortic valve |
| I080 | | | | | disorders of both mitral and aortic valves |

| | | | | | |
|---|---|---|---|---|---|
| I069 | | | | | Rheumatic aortic valve disease, unspecified |
| I068 | | | | | Other rheumatic aortic valve diseases |
| I06 | | | | | Rheumatic aortic valve diseases |
| | 3969 | | | | Diseases of the mitral and aortic valves |
| | 396 | | | | Diseases of the mitral and aortic valves |
| | 395 | | | | Disease of the aortic valve |
| | 3959 | | | | Other and unspecified diseases of aortic valve |
| | | | | K302 | revision of plastic repair of aortic valve |
| | | | | K269 | Unspecified plastic repair of aortic valve |
| | | | | K265 | aortic valve repair NEC |
| | | | | K263 | prosthetic replacement of aortic valve |
| | | | | K261 | allograft replacement of aortic valve |
| | | | | K268 | Other specified plastic repair of aortic valve |
| | | | | K264 | Replacement of aortic valve NEC |
| | | | | K262 | Xenograft replacement of aortic valve |
| | | | | K26 | Plastic repair of aortic valve |
| T820 | | | | | Mechanical complication of heart valve prosthesis |
| Z952 | | | | | Presence of prosthetic heart valve |
| Z953 | | | | | Presence of xenographic heart valve |
| | V422 | | | | Heart valve replaced by transplant |
| | V433 | | | | Replacement of heart valve (by artificial device) |
| Q238 | | | | | Other congenital malformations of aortic and mitral valves |
| | | 1065 | | | Hypertension |
| | | 1072 | | | Essential Hypertension |
| I10 | | | | | Essential (primary) hypertension |
| | | 1588 | | | Hypertrophic Cardiomyopathy |
| I421 | | | | | Obstructive hypertrophic cardiomyopathy |
| I422 | | | | | Other hypertrophic cardiomyopathy |

#### Supplementary Table 3 - Variant counts

Counts of the protein altering variants identified for each gene of interest in the UKBB exome data analysis.

i) All rare protein altering variants found in the UKBB exome data; variants used to identify genotype negative individuals:

| | | | | | | |
|---|---|---|---|---|---|---|
| *ACTC1* | *ACTN2* | *ALPK3* | *CACNA1C* | *CSRP3* | *DES* | *FHL1* |
| 39 | 403 | 947 | 675 | 123 | 225 | 104 |
| *FLNC* | *GLA* | *JPH2* | *LAMP2* | *MYBPC3* | *MYH7* | |
| 1209 | 79 | 359 | 131 | 693 | 740 | |
| *MYL2* | *MYL3* | *PLN* | *PRKAG2* | *PTPN11* | *RAF1* | |
| 86 | 84 | 21 | 200 | 160 | 208 | |
| *RIT1* | *TNNC1* | *TNNI3* | *TNNT2* | *TPM1* | *TTR* | |
| 87 | 51 | 101 | 131 | 56 | 55 | |

ii) Variants identified in the SARC-IND group:

| *ACTC1* | *MYBPC3* | *MYH7* | *MYL2* | *MYL3* | *TNNI3* | *TNNT2* | *TPM1* |
|---|---|---|---|---|---|---|---|
| 31 | 517 | 595 | 62 | 63 | 72 | 86 | 44 |

iii) Variants identified in the SARC-P/LP group:

| *MYBPC3* | *MYH7* | *TNNI3* | *TNNT2* | *TPM1* | *MYL2* | *MYL3* |
|---|---|---|---|---|---|---|
| 53 | 29 | 6 | 7 | 2 | 1 | 1 |

#### Supplementary Table 4 - manual variant curation (n=6)

| ACMG rules | Curation | hasHCM | UKBBvarID | SYMBOL | HGVSc | HGVSp | ClinVar | gnomADg_faf95_popmax | UKB_MAF |
|---|---|---|---|---|---|---|---|---|---|
| PM2, PS4_Moderate, PP1, PP3 | Likely Pathogenic | FALSE | 14:23422291:C:A | MYH7 | ENST00000355349.4:c.3134G>T | ENSP00000347507.3:p.Arg1045Leu | 42948 | 1.12E-05 | 2.49E-05 |
| PM2, PP2, BP4 | Uncertain | FALSE | 11:47332855:C:G | MYBPC3 | ENST00000545968.6:c.3449G>C | ENSP00000442795.1:p.Arg1150Thr | - | 0 | 2.49E-06 |
| PVS1, PS4, PM2, PP1, PP3 | Pathogenic | TRUE | 11:47339375:D:1 | MYBPC3 | ENST00000545968.6:c.2096del | ENSP00000442795.1:p.Pro699GlnfsTer55 | 42596 | NA | 9.97E-06 |
| PVS1, PS3, PM1, PM2, PP1, PP3 | Pathogenic | FALSE | 11:47341991:C:T | MYBPC3 | ENST00000545968.6:c.1790G>A | ENSP00000442795.1:p.Arg597Gln | 164098 | 3.15E-05 | 1.25E-05 |
| PVS1, PM2 | Likely Pathogenic | FALSE | 11:47339668:G:A | MYBPC3 | ENST00000545968.6:c.2050C>T | ENSP00000442795.1:p.Gln684Ter | - | NA | 2.49E-06 |
| PM2, PP3 | Uncertain | FALSE | 14:23416195:G:A | MYH7 | ENST00000355349.4:c.4762C>T | ENSP00000347507.3:p.Arg1588Cys | 943200 | NA | 4.98E-06 |

***Supplementary table 4:*** Manual variant curation of the six variants identified in six individuals with LVH >13mm without hypertension or valve disease. One individual had a clinical diagnosis of HCM (hasHCM). UK Biobank variant IDs (UKBBvarID) and ClinVar entry IDs (ClinVar) are shown.

#### Supplementary table 5 - Comparative lifetime risk of clinical endpoint adjusted for Sex

| Comparison | HR (95% CI) | p-value |
|---|---|---|
| **Death or MACE** | | |
| SARC-IND vs. SARC-NEG | 1.00 (0.92 to 1.08) | 0.91 |
| SARC-P/LP vs. SARC-NEG | **1.68 (1.37 to 2.06)** | **<0.001** |
| SARC-P/LP vs. SARC-IND | **1.69 (1.36 to 2.09)** | **<0.001** |
| Sex | **1.76 (1.71 to 1.81)** | **<0.001** |
| **Death** | | |
| SARC-IND vs. SARC-NEG | 1.08 (0.63 to 1.87) | 0.84 |
| SARC-P/LP vs. SARC-NEG | 0.98 (0.83 to 1.17) | 0.77 |
| SARC-P/LP vs. SARC-IND | 1.10 (0.62 to 1.95) | 0.73 |
| Sex | **1.59 (1.49 to 1.69)** | **<0.001** |
| **MACE** | | |
| SARC-IND vs. SARC-NEG | 1.00 (0.91 to 1.09) | 0.98 |
| SARC-P/LP vs. SARC-NEG | **1.84 (1.48 to 2.29)** | **<0.001** |
| SARC-P/LP vs. SARC-IND | **1.84 (1.46 to 2.33)** | **<0.001** |
| Sex | **1.81 (1.75 to 1.87)** | **<0.001** |
| **Heart Failure** | | |
| SARC-IND vs. SARC-NEG | 1.07 (0.88 to 1.29) | 0.52 |
| SARC-P/LP vs. SARC-NEG | **4.40 (3.22 to 6.02)** | **<0.001** |
| SARC-P/LP vs. SARC-IND | **4.13 (2.88 to 5.94)** | **<0.001** |
| Sex | **2.29 (2.13 to 2.47)** | **<0.001** |
| **Arrhythmia** | | |
| SARC-IND vs. SARC-NEG | 1.01 (0.91 to 1.12) | 0.83 |
| SARC-P/LP vs. SARC-NEG | **1.55 (1.18 to 2.03)** | **0.002** |
| SARC-P/LP vs. SARC-IND | **1.53 (1.14 to 2.04)** | **0.004** |
| Sex | **1.80 (1.73 to 1.87)** | **<0.001** |
| **Stroke** | | |
| SARC-IND vs. SARC-NEG | 0.88 (0.73 to 1.07) | 0.20 |
| SARC-P/LP vs. SARC-NEG | 1.53 (0.95 to 2.46) | 0.08 |
| SARC-P/LP vs. SARC-IND | **1.74 (1.04 to 2.90)** | **0.03** |
| Sex | **1.80 (1.68 to 1.92)** | **<0.001** |
| **Cardiac Arrest** | | |
| SARC-IND vs. SARC-NEG | 0.70 (0.39 to 1.23) | 0.28 |
| SARC-P/LP vs. SARC-NEG | 1.86 (0.60 to 5.79) | 0.21 |
| SARC-P/LP vs. SARC-IND | 2.67 (0.75 to 9.47) | 0.13 |
| Sex | **2.70 (2.25 to 3.26)** | **<0.001** |

***Supplementary table 5:*** Comparative lifetime risk of clinical endpoint in SARC-NEG, SARC-IND and SARC-P/LP participants. Cox proportional hazards models adjusted for sex. Major adverse cardiovascular events (MACE) consisted of heart failure, arrhythmia, stroke and cardiac arrest events. Death and MACE risk calculated using competing risk analysis from the primary clinical outcome. Results highlighted in bold reach statistical significance (P<0.05), unadjusted for multiple testing.

#### Supplementary table 6 - Comparative lifetime risk of clinical endpoint by sex

| | Female | | Male | | Sex-effect interaction | |
|---|---|---|---|---|---|---|
| | HR (95% CI) | p | HR (95% CI) | p | HR (95% CI) | p |
| **Death and MACE** | | | | | | |
| SARC-NEG vs SARC-IND | 1.09 (0.97 to 1.23) | 0.153 | 0.94 (0.85 to 1.05) | 0.272 | 1.16 (0.99 to 1.36) | 0.071 |
| SARC-NEG vs SARC-P/LP | **2.18 (1.65 to 2.89)** | **<0.001** | **1.42 (1.05 to 1.90)** | **0.02** | **1.55 (1.03 to 2.33)** | **0.04** |
| SARC-IND vs SARC-P/LP | **2.00 (1.47 to 2.71)** | **<0.001** | **1.50 (1.10 to 2.06)** | **0.01** | 1.33 (0.86 to 2.06) | 0.197 |
| **Death** | | | | | | |
| SARC-NEG vs SARC-IND | 1.07 (0.82 to 1.38) | 0.623 | 0.91 (0.72 to 1.16) | 0.457 | 1.17 (0.82 to 1.67) | 0.379 |
| SARC-NEG vs SARC-P/LP | 1.80 (0.93 to 3.46) | 0.080 | 0.64 (0.24 to 1.17) | 0.372 | 2.80 (0.86 to 9.12) | 0.087 |
| SARC-IND vs SARC-P/LP | 1.68 (0.83 to 3.39) | 0.146 | 0.70 (0.26 to 1.92) | 0.490 | 2.39 (0.70 to 8.17) | 0.164 |
| **MACE** | | | | | | |
| SARC-NEG vs SARC-IND | 1.10 (0.96 to 1.26) | 0.174 | 0.95 (0.84 to 1.07) | 0.392 | 1.16 (0.97 to 1.39) | 0.112 |
| SARC-NEG vs SARC-P/LP | **2.30 (1.68 to 3.13)** | **<0.001** | **1.61 (1.18 to 2.20)** | **0.003** | 1.43 (0.92 to 2.22) | 0.111 |
| SARC-IND vs SARC-P/LP | **2.09 (1.49 to 2.93)** | **<0.001** | **1.70 (1.22 to 2.36)** | **0.002** | 1.23 (0.77 to 1.98) | 0.383 |
| **Heart failure** | | | | | | |
| SARC-NEG vs SARC-IND | 1.18 (0.85 to 1.62) | 0.318 | 1.03 (2.49 to 5.87) | 0.822 | 1.14 (0.77 to 1.71) | 0.515 |
| SARC-NEG vs SARC-P/LP | **5.49 (3.40 to 8.87)** | **<0.001** | **3.82 (2.49 to 5.87)** | **<0.001** | 1.44 (0.76 to 2.74) | 0.268 |
| SARC-IND vs SARC-P/LP | **4.67 (2.64 to 8.25)** | **<0.001** | **3.72 (2.28 to 6.06)** | **<0.001** | 1.26 (0.59 to 2.67) | 0.547 |
| **Arrhythmia** | | | | | | |
| SARC-NEG vs SARC-IND | 1.16 (0.99 to 1.35) | 0.068 | 0.94 (0.82 to 1.08) | 0.402 | 1.23 (1.00 to 1.51) | 0.055 |
| SARC-NEG vs SARC-P/LP | **2.10 (1.44 to 3.06)** | **<0.001** | 1.32 (0.89 to 1.95) | 0.167 | 1.59 (0.92 to 2.75) | 0.094 |
| SARC-IND vs SARC-P/LP | **1.81 (1.21 to 2.72)** | **0.004** | 1.40 (0.92 to 2.12) | 0.112 | 1.30 (0.72 to 2.32) | 0.382 |
| **Stroke** | | | | | | |
| SARC-NEG vs SARC-IND | 0.89 (0.69 to 1.14) | 0.344 | 0.89 (0.69 to 1.14) | 0.344 | 0.94 (0.33 to 2.65) | 0.864 |
| SARC-NEG vs SARC-P/LP | 1.45 (0.75 to 2.79) | 0.266 | 1.45 (0.75 to 2.79) | 0.266 | 0.97 (0.65 to 1.44) | 0.910 |
| SARC-IND vs SARC-P/LP | 1.59 (0.68 to 3.76) | 0.287 | 1.64 (0.81 to 3.29) | 0.167 | 0.98 (0.32 to 2.95) | 0.965 |
| **Cardiac arrest** | | | | | | |
| SARC-NEG vs SARC-IND | 0.39 (0.10 to 1.57) | 0.185 | 0.85 (0.45 to 1.59) | 0.6104 | 0.46 (0.10 to 2.12) | 0.318 |
| SARC-NEG vs SARC-P/LP | - | - | 2.92 (0.94 to 9.08) | 0.0649 | - | - |
| SARC-IND vs SARC-P/LP | - | - | 3.43 (0.94 to 12.5) | 0.061 | - | - |

***Supplementary table 6 -*** Comparative lifetime risk of clinical endpoint in SARC-NEG, SARC-IND and SARC-P/LP participants stratified by sex. Major adverse cardiovascular events (MACE) consisted of heart failure, arrhythmia, stroke and cardiac arrest events. Death and MACE risk calculated using competing risk analysis from the primary clinical outcome. Sex-effect interaction calculated using cox proportional hazard model with inclusion of a sexgenotype interaction term, with HR representing female risk compared with male. Results highlighted in bold reach statistical significance ($p<0.05$), unadjusted for multiple testing.

#### Supplementary table 7 – Incident risk of clinical events

| | HR (95% CI) | p |
|---|---|---|
| **Death or MACE** | | |
| SARC-NEG vs SARC-IND | 1.03 (0.93 to 1.14) | 0.561 |
| SARC-NEG vs SARC-P/LP | **1.54 (1.18 to 2.01)** | **0.001** |
| SARC-IND vs SARC-P/LP | **1.50 (1.13 to 1.98)** | **0.005** |
| **Death** | | |
| SARC-NEG vs SARC-IND | 0.98 (0.82 to 1.17) | 0.809 |
| SARC-NEG vs SARC-P/LP | 1.10 (0.64 to 1.90) | 0.723 |
| SARC-IND vs SARC-P/LP | 1.13 (0.64 to 1.99) | 0.680 |
| **MACE** | | |
| SARC-NEG vs SARC-IND | 1.05 (0.94 to 1.19) | 0.378 |
| SARC-NEG vs SARC-P/LP | **1.76 (1.30 to 2.38)** | **<0.001** |
| SARC-IND vs SARC-P/LP | **1.67 (1.21 to 2.30)** | **0.002** |
| **Heart failure** | | |
| SARC-NEG vs SARC-IND | 0.96 (0.76 to 1.22) | 0.728 |
| SARC-NEG vs SARC-P/LP | **3.23 (2.10 to 4.97)** | **<0.001** |
| SARC-IND vs SARC-P/LP | **3.37 (2.07 to 5.49)** | **<0.001** |
| **Stroke** | | |
| SARC-NEG vs SARC-IND | 1.07 (0.81 to 1.41) | 0.635 |
| SARC-NEG vs SARC-P/LP | **1.97 (1.02 to 3.79)** | **0.04** |
| SARC-IND vs SARC-P/LP | 1.84 (0.91 to 3.73) | 0.09 |
| **Arrhythmia** | | |
| SARC-NEG vs SARC-IND | 1.06 (0.93 to 1.22) | 0.354 |
| SARC-NEG vs SARC-P/LP | **1.58 (1.11 to 2.24)** | **0.01** |
| SARC-IND vs SARC-P/LP | **1.48 (1.02 to 2.15)** | **0.04** |
| **Cardiac arrest** | | |
| SARC-NEG vs SARC-IND | 0.68 (0.36 to 1.28) | 0.15 |
| SARC-NEG vs SARC-P/LP | 2.31 (0.74 to 7.18) | 0.2 |
| SARC-IND vs SARC-P/LP | 3.39 (0.93 to 12.3) | 0.064 |

***Supplementary table 7*** - Incident risk of clinical events in SARC-NEG, SARC-IND and SARC-P/LP participants. Cox proportional hazards models adjusted for age at recruitment and sex. Major adverse cardiovascular events (MACE) defined as heart failure, arrhythmia, stroke or cardiac arrest events. Death and MACE calculated using competing risk analysis from primary clinical outcome. Results highlighted in bold reach statistical significance ($p<0.05$), unadjusted for multiple testing.

#### Supplementary Table 8 – Sensitivity analysis excluding subjects with cardiomyopathy and/or cardiomyopathy from the endpoint

A) Sensitivity analysis excluding cardiomyopathy from outcome

| | HR (95% CI) | p |
|---|---|---|
| Death and MACE (excluding cardiomyopathy) | | |
| SARC-NEG vs SARC-IND | 0.98 (0.89 to 1.07) | 0.611 |
| SARC-NEG vs SARC-P/LP | **1.53 (1.19 to 1.96)** | **<0.001** |
| SARC-IND vs SARC-P/LP | **1.57 (1.20 to 2.04)** | **<0.001** |
| MACE (excluding cardiomyopathy) | | |
| SARC-NEG vs SARC-IND | 0.98 (0.89 to 1.07) | 0.627 |
| SARC-NEG vs SARC-P/LP | **1.49 (1.18 to 1.89)** | **<0.001** |
| SARC-IND vs SARC-P/LP | **1.53 (1.18 to 1.97)** | **0.001** |
| Heart failure (excluding cardiomyopathy) | | |
| SARC-NEG vs SARC-IND | 0.98 (0.80 to 1.21) | 0.857 |
| SARC-NEG vs SARC-P/LP | **1.88 (1.15 to 3.07)** | **0.01** |
| SARC-IND vs SARC-P/LP | **1.92 (1.13 to 3.26)** | **0.02** |

B) Sensitivity analysis excluding participants with a diagnostic label of cardiomyopathy

| | HR (95% CI) | p |
|---|---|---|
| Death and MACE (excluding cardiomyopathy) | | |
| SARC-NEG vs SARC-IND | 0.99 (0.91 to 1.07) | 0.82 |
| SARC-NEG vs SARC-P/LP | **1.31 (1.03 to 1.67)** | **0.026** |
| SARC-IND vs SARC-P/LP | **1.33 (1.03 to 1.71)** | **0.028** |
| MACE (excluding cardiomyopathy) | | |
| SARC-NEG vs SARC-IND | 0.99 (0.90 to 1.08) | 0.80 |
| SARC-NEG vs SARC-P/LP | **1.34 (1.03 to 1.75)** | **0.032** |
| SARC-IND vs SARC-P/LP | **1.36 (1.02 to 1.80)** | **0.034** |
| Heart failure (excluding cardiomyopathy) | | |
| SARC-NEG vs SARC-IND | 1.00 (0.80 to 1.25) | 0.99 |
| SARC-NEG vs SARC-P/LP | **1.78 (1.01 to 3.14)** | **0.047** |
| SARC-IND vs SARC-P/LP | 1.78 (0.97 to 3.27) | 0.062 |

***Supplementary table 8*** - Sensitivity analysis excluding cardiomyopathy from lifetime risk of outcomes (A) and excluding participants with any diagnostic label of cardiomyopathy (n=671). Cox proportional hazards models adjusted for sex. Major adverse cardiovascular events (MACE) defined as heart failure (excluding cardiomyopathy), arrhythmia, stroke or cardiac arrest events. Death and MACE calculated using competing risk analysis from primary clinical outcome. Results highlighted in bold reach statistical significance ($P<0.05$), unadjusted for multiple testing.

#### Supplementary Table 9 – Clinical outcomes stratified by LV wall thickness

| | Adjusted for BP and sex | |
|---|---|---|
| | **HR (95% CI)** | **p** |
| **Death* or MACE** | | |
| 13-15 vs <13mm | **1.69 (1.24 to 2.30)** | **<0.001** |
| >15 vs <13mm | **3.00 (1.86 to 4.86)** | **<0.001** |
| >15 vs 13-15 | **1.78 (1.02 to 3.12)** | **0.04** |
| Per mm increase | **1.15 (1.11 to 1.20)** | **<0.001** |
| **MACE** | | |
| 13-15 vs <13mm | **1.79 (1.31 to 2.43)** | **<0.001** |
| >15 vs <13mm | **3.16 (1.95 to 5.11)** | **<0.001** |
| >15 vs 13-15 | **1.77 (1.01 to 3.10)** | **0.05** |
| Per mm increase | **1.17 (1.12 to 1.21)** | **<0.001** |
| **Heart failure** | | |
| 13-15 vs <13mm | **2.53 (1.22 to 5.24)** | **0.01** |
| >15 vs <13mm | **8.26 (3.60 to 18.9)** | **<0.001** |
| >15 vs 13-15 | **3.27 (1.13 to 9.42)** | **0.01** |
| Per mm increase | **1.36 (1.25 to 1.49)** | **<0.001** |
| **Arrhythmia** | | |
| 13-15 vs <13mm | **1.93 (1.36 to 2,73)** | **<0.001** |
| >15 vs <13mm | **2.42 (1.29 to 4.53)** | **0.006** |
| >15 vs 13-15 | 1.25 (0.62 to 2.54) | 0.528 |
| Per mm increase | **1.14 (1.09 to 1.19)** | **<0.001** |
| **Stroke** | | |
| 13-15 vs <13mm | 1.03 (0.45 to 2.33) | 0.948 |
| >15 vs <13mm | 2.19 (0.70 to 6.86) | 0.179 |
| >15 vs 13-15 | 2.13 (0.53 to 8.52) | 0.285 |
| Per mm increase | **1.15 (1.05 to 1.25)** | **0.001** |

***Supplementary table 9*** - Lifetime risk of clinical events stratified by LV wall thickness (<13mm, 13 to 15mm and >15mm) in individuals with imaging and genetic data (N=39,559). Cox proportional hazards model adjusted for systolic blood pressure and sex. Major adverse cardiovascular events (MACE) defined as heart failure, arrhythmia, stroke or cardiac arrest events. Death and MACE calculated using competing risk analysis from primary clinical outcome. *One death reported in 13-15mm group and zero deaths reported in >15mm group. Results highlighted in bold reach statistical significance (P<0.05), unadjusted for multiple testing.

#### Supplementary table 10 - Clinical outcomes in individuals with imaging and genetic data stratified by LV wall thickness

| | Unadjusted for genotype | | Adjusted for genotype | |
|---|---|---|---|---|
| | **HR (95% CI)** | **p** | **HR (95% CI)** | **p** |
| **Death* and MACE** | | | | |
| 13-15 vs <13mm | **1.68 (1.24 to 2.26)** | **<0.001** | **1.65 (1.23 to 2.23)** | **<0.001** |
| >15 vs <13mm | **2.33 (1.35 to 4.04)** | **0.002** | **2.28 (1.32 to 3.96)** | **0.003** |
| >15 vs 13-15 | 1.37 (0.74 to 2.54) | 0.294 | 1.37 (0.74 to 2.54) | 0.303 |
| Per mm increase | **1.13 (1.09 to 1.18)** | **<0.001** | **1.13 (1.09 to 1.17)** | **<0.001** |
| **MACE** | | | | |
| 13-15 vs <13mm | **1.75 (1.30 to 2.36)** | **<0.001** | **1.72 (1.28 to 2.32)** | **<0.001** |
| >15 vs <13mm | **2.43 (1.40 to 4.20)** | **0.002** | **2.37 (1.37 to 4.11)** | **0.002** |
| >15 vs 13-15 | 1.39 (0.75 to 2.57) | 0.296 | 1.38 (0.74 to 2.55) | 0.306 |
| Per mm increase | **1.14 (1.10 to 1.19)** | **<0.001** | **1.14 (1.09 to 1.18)** | **<0.001** |
| **Heart failure** | | | | |
| 13-15 vs <13mm | **2.66 (1.34 to 5.31)** | **0.005** | **2.39 (1.19 to 4.79)** | **0.01** |
| >15 vs <13mm | **7.23 (2.93 to 17.8)** | **<0.001** | **6.43 (2.59 to 16.0)** | **<0.001** |
| >15 vs 13-15 | 2.71 (0.91 to 8.10) | 0.073 | 2.69 (0.90 to 8.02) | 0.076 |
| Per mm increase | **1.34 (1.22 to 1.48)** | **<0.001** | **1.32 (1.20 to 1.45)** | **<0.001** |
| **Arrhythmia** | | | | |
| 13-15 vs <13mm | **1.81 (1.29 to 2.54)** | **<0.001** | **1.80 (1.28 to 2.52)** | **<0.001** |
| >15 vs <13mm | 1.18 (0.49 to 2.84) | 0.717 | 1.16 (0.48 to 2.80) | 0.740 |
| >15 vs 13-15 | 0.65 (0.26 to 1.66) | 0.366 | 0.65 (0.25 to 1.65) | 0.359 |
| Per mm increase | **1.11 (1.06 to 1.16)** | **<0.001** | **1.11 (1.06 to 1.16)** | **<0.001** |
| **Stroke** | | | | |
| 13-15 vs <13mm | 1.54 (0.79 to 3.02) | 0.209 | 1.54 (0.78 to 3.03) | 0.209 |
| >15 vs <13mm | **3.31 (1.22 to 8.93)** | **0.02** | **3.30 (1.22 to 8.93)** | **0.02** |
| >15 vs 13-15 | 2.14 (0.66 to 6.96) | 0.204 | 2.14 (0.66 to 6.96) | 0.205 |
| Per mm increase | **1.15 (1.06 to 1.25)** | **<0.001** | **1.15 (1.06 to 1.26)** | **<0.001** |

***Supplementary table 10*** - Lifetime risk of clinical events stratified by LV wall thickness (<13mm, 13 to 15mm and >15mm) in individuals with imaging and genetic data (n=17,447). Cox proportional hazards model adjusted for systolic blood pressure and sex. Major adverse cardiovascular events (MACE) defined as heart failure, arrhythmia, stroke or cardiac arrest events. Death and MACE calculated using competing risk analysis from primary clinical outcome. *One death reported in 13-15mm group and zero deaths reported in >15mm group. Results highlighted in bold reach statistical significance (P<0.05), unadjusted for multiple testing.

#### Supplementary Table 11 - Clinical outcomes stratified by LV fractal dimension

| | Model 1 | | Model 2 | |
|---|---|---|---|---|
| | **HR (95% CI)** | **p** | **HR (95% CI)** | **p** |
| **Mean global FD** | | | | |
| Per quantile | **1.31 (1.12 to 1.54)** | **0.001** | 1.03 (0.88 to 1.22) | 0.68 |
| Per 0.1 unit increase | **1.04 (1.01 to 1.08)** | **0.01** | 1.00 (0.97 to 1.04) | 0.97 |
| **Mean apical FD** | | | | |
| Per quantile | **1.21 (1.08 to 1.35)** | **<0.001** | 1.09 (0.97 to 1.22) | 0.16 |
| Per 0.1 unit increase | **1.05 (1.02 to 1.09)** | **0.002** | 1.02 (0.99 to 1.06) | 0.18 |
| **Mean basal FD** | | | | |
| Per quantile | 1.15 (1.01 to 1.32) | 0.04 | 0.95 (0.83 to 1.09) | 0.49 |
| Per 0.1 unit increase | 1.02 (0.98 to 1.05) | 0.32 | 0.98 (0.94 to 1.01) | 0.15 |

***Supplementary table 11 -*** Lifetime risk of clinical events in individuals with fractal dimension measurements. Cox proportional hazards model 1 adjusted for systolic blood pressure, sex and LV wall thickness. Model 2 as model 1 with additional co-variate of LA volume. Major adverse cardiovascular events (MACE) defined as heart failure, arrhythmia, stroke or cardiac arrest events. Death and MACE calculated using competing risk analysis from primary clinical outcome. Results highlighted in bold reach statistical significance ($p<0.05$), unadjusted for multiple testing.

#### Supplementary table 12 - Clinical outcomes in individuals with imaging and genetic data stratified by genotype and adjusted for LV wall thickness

| | Unadjusted for LV max wall thickness | | Adjusted for LV max wall thickness | |
|---|---|---|---|---|
| | HR (95% CI) | p | HR (95% CI) | p |
| **Death and MACE** | | | | |
| SARC-NEG vs SARC-IND | 1.20 (0.89 to 1.61) | 0.236 | 1.17 (0.87 to 1.57) | 0.306 |
| SARC-NEG vs SARC-P/LP | **2.53 (1.20 to 5.32)** | **0.01** | 1.99 (0.95 to 4.21) | 0.070 |
| SARC-IND vs SARC-P/LP | 2.11 (0.95 to 4.69) | 0.0653 | 1.71 (0.77 to 3.80) | 0.190 |
| **Death** | | | | |
| SARC-NEG vs SARC-IND | 0.87 (0.12 to 6.34) | 0.89086 | 0.87 (0.12 to 6.37) | 0.894 |
| SARC-NEG vs SARC-P/LP | - | - | - | - |
| SARC-IND vs SARC-P/LP | - | - | - | - |
| **MACE** | | | | |
| SARC-NEG vs SARC-IND | **1.75 (1.30 to 2.36)** | **<0.001** | **1.72 (1.28 to 2.32)** | **<0.001** |
| SARC-NEG vs SARC-P/LP | **2.43 (1.40 to 4.20)** | **0.002** | **2.37 (1.37 to 4.11)** | **0.002** |
| SARC-IND vs SARC-P/LP | 2.15 (0.97 to 4.78) | 0.059 | 1.72 (0.77 to 3.83) | 0.183 |
| **Heart failure** | | | | |
| SARC-NEG vs SARC-IND | **2.48 (1.30 to 4.71)** | **0.006** | **2.35 (1.23 to 4.48)** | **0.009** |
| SARC-NEG vs SARC-P/LP | **13.9 (5.15 to 37.7)** | **<0.001** | **8.05 (2.91 to 22.3)** | **<0.001** |
| SARC-IND vs SARC-P/LP | **5.63 (1.76 to 18. 0)** | **0.004** | **3.42 (1.06 to 11.1)** | **0.04** |
| **Arrhythmia** | | | | |
| SARC-NEG vs SARC-IND | 1.12 (0.78 to 1.60) | 0.535 | 1.10 (0.77 to 1.57) | 0.619 |
| SARC-NEG vs SARC-P/LP | 1.37 (0.44 to 4.26) | 0.585 | 1.10 (0.35 to 3.43) | 0.871 |
| SARC-IND vs SARC-P/LP | 1.22 (0.37 to 4.01) | 0.738 | 1.00 (0.31 to 3.29) | 0.995 |
| **Stroke** | | | | |
| SARC-NEG vs SARC-IND | 0.92 (0.44 to 1.96) | 0.837 | 0.89 (0.42 to 1.89) | 0.766 |
| SARC-NEG vs SARC-P/LP | 1.62 (0.23 to 11.5) | 0.632 | 1.19 (0.17 to 8.55) | 0.863 |
| SARC-IND vs SARC-P/LP | 1.75 (0.22 to 14.2) | 0.601 | 1.33 (0.16 to 10.9) | 0.788 |

***Supplementary table 12*** - Lifetime risk of clinical events in SARC-NEG, SARC-IND and SARC-P/LP participants with imaging and genomic data (n=17,447). Cox proportional hazards model adjusted for LV max wall thickness and/or sex. Major adverse cardiovascular events (MACE) defined as heart failure, arrhythmia, stroke or cardiac arrest events. Death and MACE calculated using competing risk analysis from primary clinical outcome. No death events in participants with SARC-P/LP. Results highlighted in bold reach statistical significance (P<0.05), unadjusted for multiple testing.

#### Supplementary table 13 - Clinical outcomes stratified by genotype and adjusted for LV wall thickness and LA volume

| | Unadjusted for LV max wall thickness or LA max volume | | Adjusted for LV max wall thickness and LA max volume | |
|---|---|---|---|---|
| | **HR (95% CI)** | **p** | **HR (95% CI)** | **p** |
| **Death and MACE** | | | | |
| SARC-NEG vs SARC-IND | 1.20 (0.89 to 1.61) | 0.23 | 1.10 (0.81 to 1.49) | 0.54 |
| SARC-NEG vs SARC-P/LP | **2.53 (1.21 to 5.33)** | **0.01** | 1.50 (0.67 to 3.37) | 0.32 |
| SARC-IND vs SARC-P/LP | 2.11 (0.95 to 4.69) | 0.07 | 1.37 (0.58 to 3.22) | 0.47 |
| LV max wall thickness (per mm increase) | - | - | **1.09 (1.05 to 1.13)** | **<0.001** |
| LA max volume (per ml increase) | - | - | **1.02 (1.01 to 1.02)** | **<0.001** |
| **Death** | | | | |
| SARC-NEG vs SARC-IND | 0.85 (0.12 to 6.23) | 0.88 | 0.95 (0.13 to 6.90) | 0.96 |
| SARC-NEG vs SARC-P/LP | **-** | **-** | - | - |
| SARC-IND vs SARC-P/LP | **-** | **-** | - | - |
| LV max wall thickness (per mm increase) | **-** | **-** | 0.93 (0.73 to 1.18) | 0.56 |
| LA max volume (per ml increase) | **-** | **-** | 1.00 (0.98 to 1.01) | 0.60 |
| **MACE** | | | | |
| SARC-NEG vs SARC-IND | 1.21 (0.89 to 1.63) | 0.22 | 1.10 (0.81 to 1.49) | 0.54 |
| SARC-NEG vs SARC-P/LP | **2.60 (1.24 to 5.48)** | **0.01** | 1.52 (0.68 to 3.42) | 0.31 |
| SARC-IND vs SARC-P/LP | 2.15 (0.97 to 4.78) | 0.06 | 1.39 (0.59 to 3.27) | 0.46 |
| LV max wall thickness (per mm increase) | - | - | **1.10 (1.05 to 1.14)** | **<0.001** |
| LA max volume (per ml increase) | - | - | **1.02 (1.01 to 1.02)** | **<0.001** |
| **Heart failure** | | | | |
| SARC-NEG vs SARC-IND | **2.48 (1.30 to 4.71)** | **0.006** | **2.21 (1.16 to 4.23)** | **0.006** |
| SARC-NEG vs SARC-P/LP | **13.9 (5.15 to 37.7)** | **<0.001** | **5.25 (1.62 to 17.0)** | **0.02** |
| SARC-IND vs SARC-P/LP | **5.63 (1.76 to 18. 0)** | **0.004** | 2.38 (0.64 to 8.87) | 0.20 |
| LV max wall thickness (per mm increase) | **-** | **-** | **1.23 (1.11 to 1.35)** | **<0.001** |
| LA max volume (per ml increase) | - | - | **1.02 (1.01 to 1.02)** | **<0.001** |
| **Arrhythmia** | | | | |
| SARC-NEG vs SARC-IND | 1.12 (0.78 to 1.60) | **0.54** | 0.95 (0.66 to 1.37) | **0.80** |
| SARC-NEG vs SARC-P/LP | 1.37 (0.44 to 4.26) | **0.59** | 0.67 (0.17 to 2.69) | **0.57** |
| SARC-IND vs SARC-P/LP | 1.22 (0.37 to 4.01) | **0.74** | 0.70 (0.17 to 2.94) | **0.63** |

| | | | | |
|---|---|---|---|---|
| LV max wall thickness (per mm increase) | **-** | **-** | **1.05 (1.00 to 1.10)** | **0.04** |
| LA max volume (per ml increase) | **-** | **-** | **1.02 (1.02 to 1.02)** | **<0.001** |
| **Stroke** | | | | |
| SARC-NEG vs SARC-IND | 0.92 (0.44 to 1.96) | 0.84 | 1.28 (0.57 to 2.87) | 0.56 |
| SARC-NEG vs SARC-P/LP | 1.62 (0.23 to 11.5) | 0.63 | 1.56 (0.19 to 13.0) | 0.68 |
| SARC-IND vs SARC-P/LP | 1.75 (0.22 to 14.2) | 0.60 | 1.22 (0.17 to 8.78) | 0.84 |
| LV max wall thickness (per mm increase) | - | - | **1.16 (1.07 to 1.26)** | **<0.001** |
| LA max volume (per ml increase) | - | - | 1.00 (1.00 to 1.01) | 0.55 |

***Supplementary table 13*** - Lifetime risk of clinical events in SARC-NEG, SARC-IND and SARC-P/LP participants with imaging data (N=39,274). Cox proportional hazards model adjusted for LV wall thickness and LA volumes and/or sex. Major adverse cardiovascular events (MACE) defined as heart failure, arrhythmia, stroke or cardiac arrest events. Death and MACE calculated using competing risk analysis from primary clinical outcome. No death events in participants with SARC-P/LP. Results highlighted in bold reach statistical significance ($p<0.05$), unadjusted for multiple testing.

#### Supplementary Figure 1 - Cumulative hazard curves for lifetime risk of primary clinical endpoint stratified by genotype in males and females

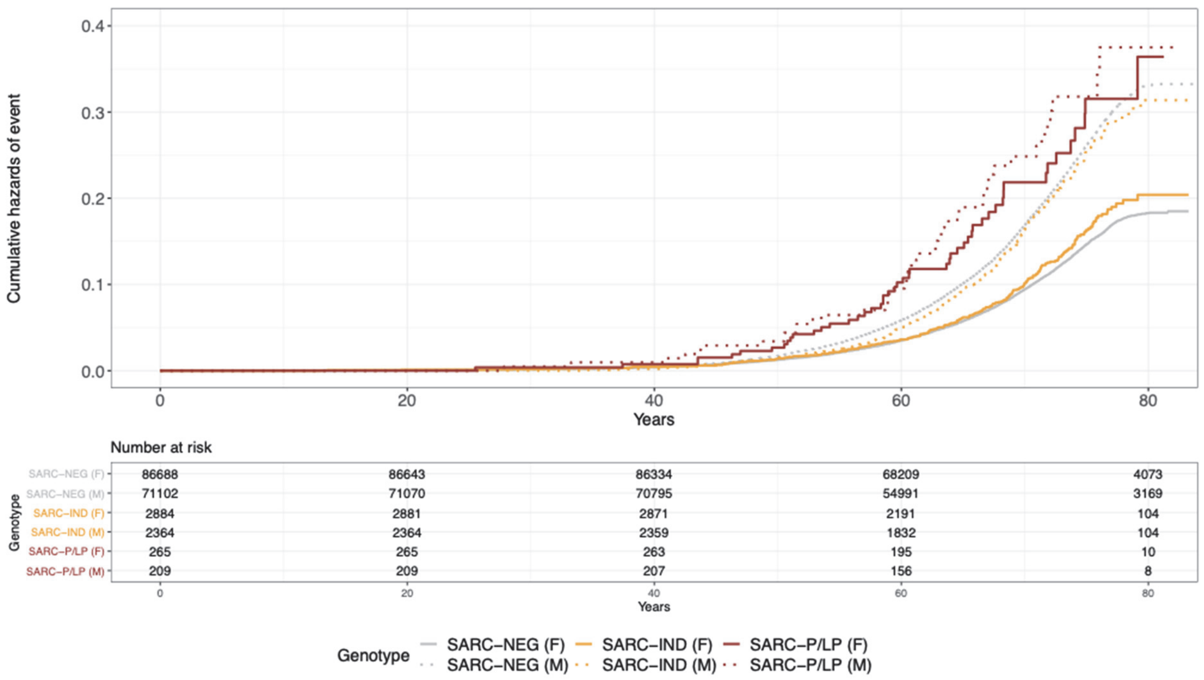

Cumulative hazard curves for lifetime risk of death or major adverse cardiovascular events stratified by genotype in males (solid line) and females (dotted line). MACE defined as heart failure, arrhythmia, stroke and cardiac arrest events.

#### Supplementary Figure 2 - Cumulative hazard curves for lifetime risk of clinical events stratified by LV maximum wall thickness

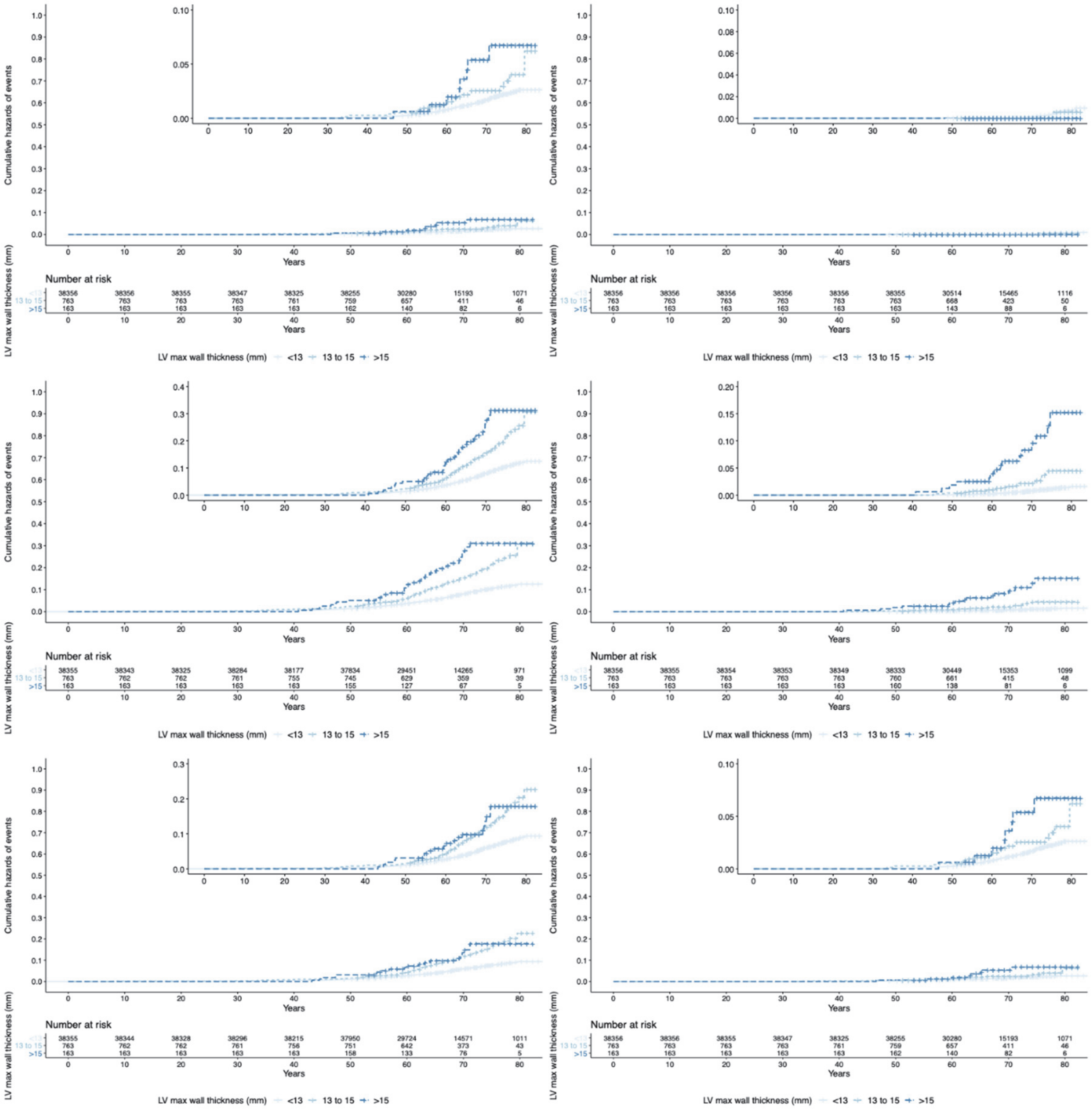

Cumulative hazard curves for lifetime risk of clinical events stratified by LV maximum wall thickness (<13mm, 13 to 15mm and >15mm) in individuals with available imaging data (N=39,274). (Top panel left-right) Death and major adverse cardiovascular events (MACE), death, (middle panel left-right) MACE, heart failure, (bottom panel left-right) arrhythmia, and stroke events.  MACE defined as heart failure, arrhythmia, stroke and cardiac arrest events.

#### Supplementary Figure 3 - Cumulative hazard curves for lifetime risk of clinical events stratified by LV maximum wall thickness in participants with both imaging and genetic data

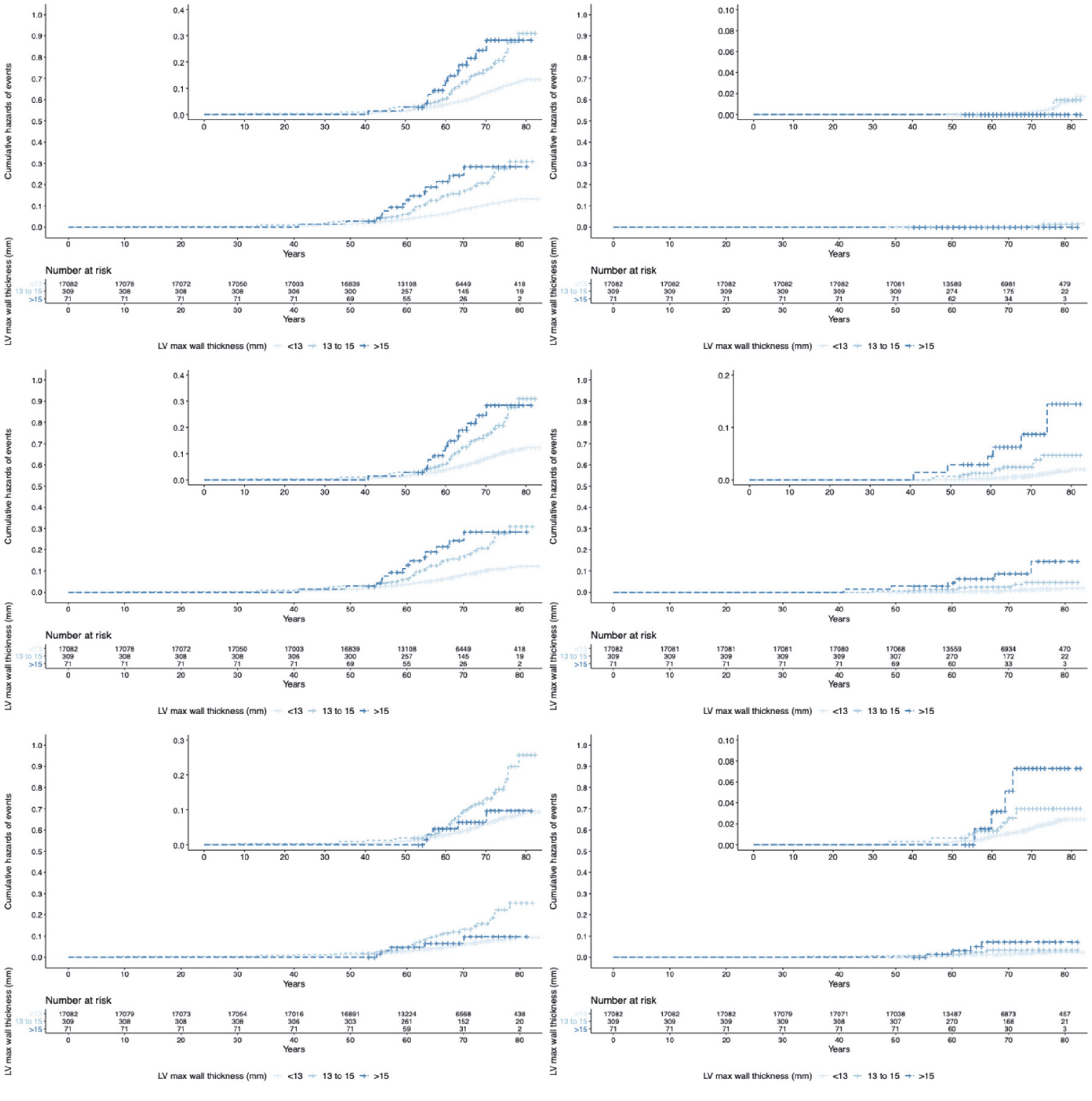

Cumulative hazard curves for lifetime risk of clinical events stratified by LV maximum wall thickness (<13mm, 13 to 15mm and >15mm) in individuals with available imaging and genetic data (N=17,447). (Top panel left-right) Death and major adverse cardiovascular events (MACE), death, (middle panel left-right) MACE, heart failure, (bottom panel left-right) arrhythmia, and stroke events. MACE defined as heart failure, arrhythmia, stroke and cardiac arrest events.

### Supplementary Figure 4 - Cumulative hazard curves for lifetime risk of clinical events stratified by quintiles of LV fractal dimensions

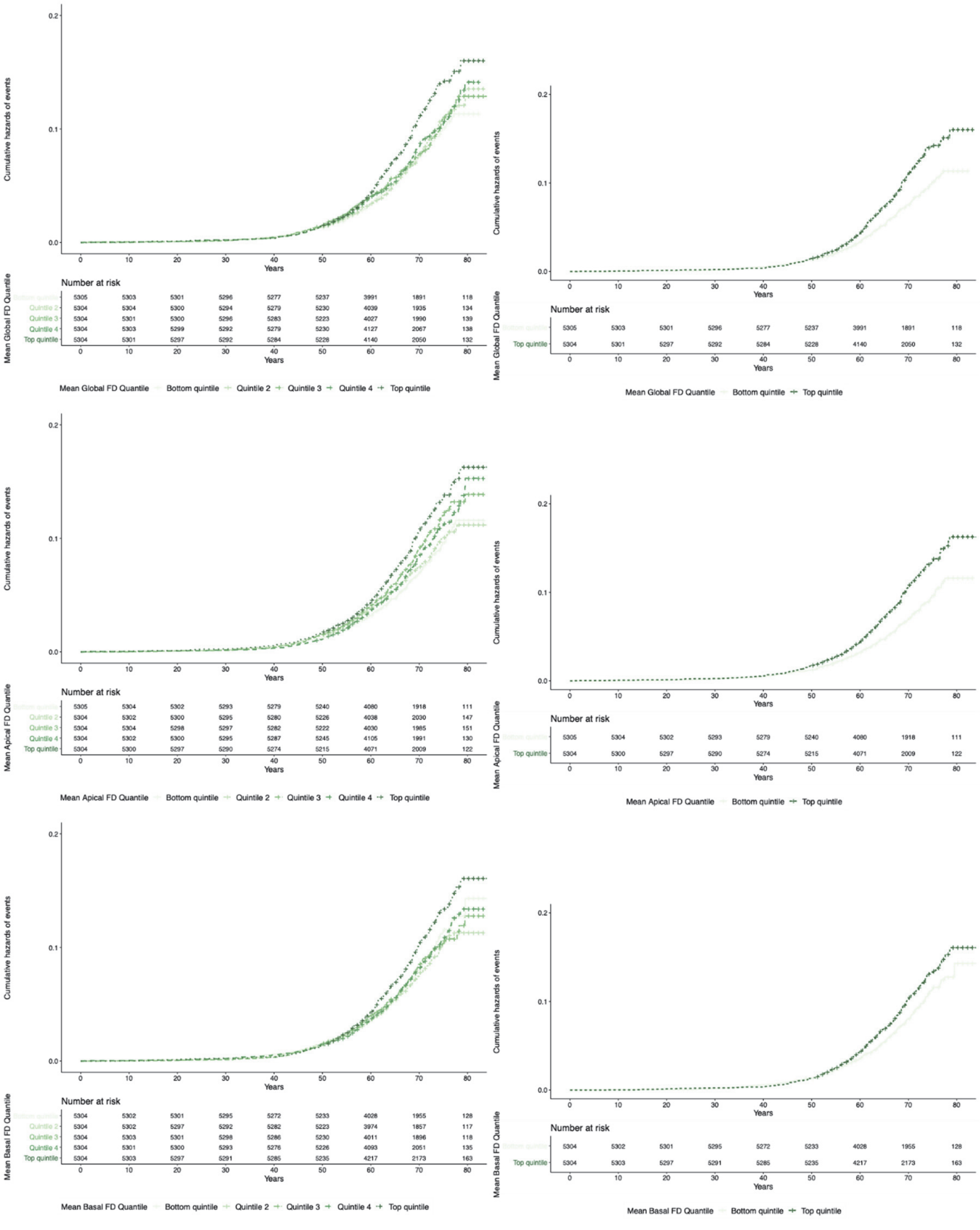

Cumulative hazard curves for lifetime risk of death or major adverse cardiovascular events stratified by quintiles of LV fractal dimensions (FD) in individuals. (Top panel left-right) mean global FD for all quintiles, and top and bottom quintiles, (middle panel left-right) mean apical FD for all quintiles, and top and bottom quintiles, and (bottom panel left-right) mean basal FD for all quintiles, and top and bottom quintiles.  MACE defined as heart failure, arrhythmia, stroke and cardiac arrest events.

#### Supplementary Figure 5 - Cumulative hazard curves for incident clinical outcomes stratified by genotypes

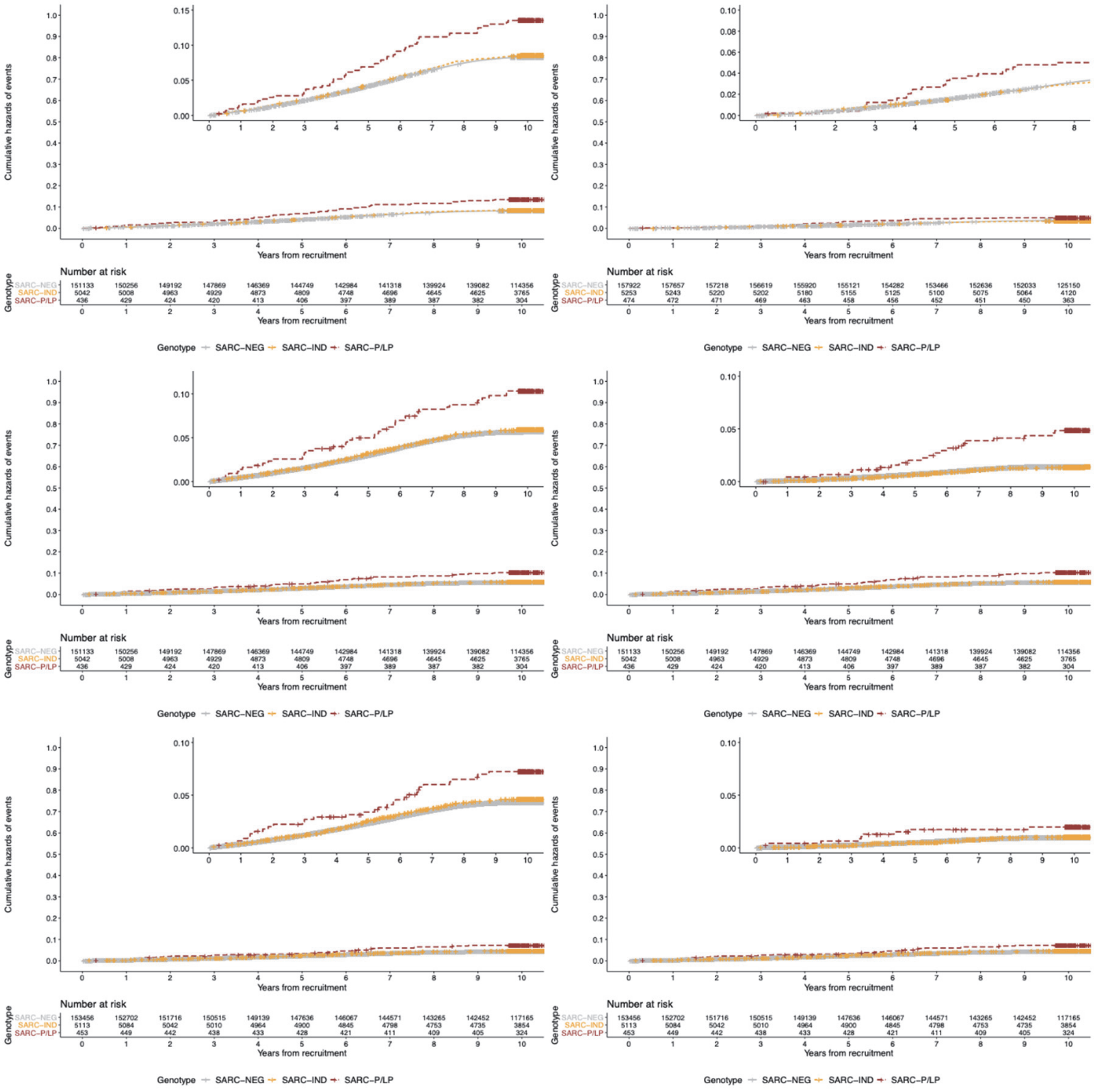

Cumulative hazard curve for incident clinical outcomes stratified by genotypes: individuals without sarcomeric variants (SARC-NEG), sarcomeric variants of indeterminate significance (SARC-IND) and P/LP sarcomeric variants (SARC-P/LP). (Top panels, left-right) Death or major adverse cardiovascular events (MACE), death, (middle panels, left-right) MACE, heart failure, (bottom panels, left-right) arrhythmia, and stroke. MACE defined as heart failure, arrhythmia, stroke or cardiac arrest events.

#### SARC-IND Sub-group analysis

A secondary analysis explored whether variants in the SARC-IND category might be further stratified based on features suggested as important but not yet universally adopted in diagnostic practice. We explore several approaches to stratify variants in the SARC-IND group, and to look at additional genes and variant classes that were removed from the analysis where an association with HCM is not robustly established (A-F, below). For all groups, we compared CMR 2D phenotypes to the SARC-NEG individuals, and conducted primary clinical outcome analyses. The groups of SARC-IND variants investigated are as follows:

A) missense variants in the SARC-IND group found in amino acid residues previously reported to have a high prior likelihood of pathogenicity in HCM[13]: TPM1 (whole gene), MYH7 (residues 167-969), MYBPC3 (178-311, 432-597, 750-858, 1194-1266), TNNI3 (136-171, 186-209), TNNT2 (79-104, 151-179, 278-286) and MYL3 (149-156).

B) all other variant types in the SARC-IND group such as truncating variants that are predicted to escape NMD, indels, and loss of start and stop codons.

C) ClinGen "moderate evidence" genes: we evaluated the clinical phenotype associated with variants in genes with moderate evidence of involvement in HCM (*CSRP3* (n=107 variants), *TNNC1* (n=46 variants), and *JPH2* (n=309 variants)), which were previously excluded from the SARC-IND group.

D) we investigated truncating variants in the 7 sarcomeric genes where missense variants are causative of HCM, but where haploinsufficiency or protein-truncation are not robustly established mechanisms of disease *(MYH7, MYL2, MYL3, ACTC1, TNNI3, TNNT2, TPM1*).

E) we assessed individuals with compound heterozygous or homozygous SARC-IND variants.

All groups here ((A) to (E)) did not reveal significant differences (p>0.05). On assessment of the groups, aggregation of groups (A) and (B), denoted here as "SARC-IND high probability", revealed an increased risk of the primary clinical outcome in older age at landmark survival analysis conducted on individuals free from the primary clinical outcome at ages 40, 50, 60 and 70 years old (from age 60 years old: HR 1.23, 95% CI 1.03-1.46, p=0.02; from age 70 years old: HR 1.48, 95% CI 1.14-1.94, p=0.004) (**Table S14; Figure S6**). This was not found when the groups were assessed independently. Group (B) variants, deemed “SARC-IND low probability”, were assessed alongside the aggregate of groups (A) and (C) (**Table S14; Figure S6**).

#### Supplementary Table 14 - Landmark analysis of clinical outcomes stratified by genotype SARC-IND high probability and SARC-IND low probability

| | HR (95% CI) | p-value |
|---|---|---|
| **Lifetime probability (vs. SARC-NEG)** | | |
| SARC-IND low probability | 0.96 (0.88 to 1.06) | 0.43 |
| SARC-IND high probability | 1.12 (0.96 to 1.29) | 0.14 |
| SARC-P/LP | 1.74 (1.42 to 2.13) | <0.001 |
| **From age 40 (vs. SARC-NEG)** | | |
| SARC-IND low probability | 0.97 (0.88 to 1.06) | 0.5 |
| SARC-IND high probability | 1.13 (0.98 to 1.31) | 0.10 |
| SARC-P/LP | 1.73 (1.40 to 2.13) | <0.001 |
| **From age 50 (vs. SARC-NEG)** | | |
| SARC-IND low probability | 0.96 (0.87 to 1.06) | 0.4 |
| SARC-IND high probability | 1.14 (0.97 to 1.33) | 0.11 |
| SARC-P/LP | 1.69 (1.35 to 2.11) | <0.001 |
| **From age 60 (vs. SARC-NEG)** | | |
| SARC-IND low probability | 0.98 (0.88 to 1.10) | 0.8 |
| SARC-IND high probability | 1.23 (1.03 to 1.46) | 0.021 |
| SARC-P/LP | 1.48 (1.13 to 1.95) | 0.005 |
| **From age 70 (vs. SARC-NEG)** | | |
| SARC-IND low probability | 0.91 (0.75 to 1.11) | 0.4 |
| SARC-IND high probability | 1.48 (1.14 to 1.94) | 0.004 |
| SARC-P/LP | 1.16 (0.70 to 1.93) | 0.6 |

***Supplementary Table 14*** - Landmark analysis of clinical events in genotyped individuals with SARC-IND separated into those with high probability and low probability variants. Cox proportional hazards model adjusted for age at recruitment and sex. Major adverse cardiovascular events (MACE) defined as heart failure, arrhythmia, stroke or cardiac arrest events. Death and MACE calculated using competing risk analysis from primary clinical outcome. Individuals with events preceding the age from landmark analysis were excluded. Results highlighted in bold reach statistical significance ($p<0.05$), unadjusted for multiple testing.

#### Supplementary Figure 6 - Cumulative hazard curves of landmark analysis for death or MACE stratified by SARC-IND high probability and low probability variants

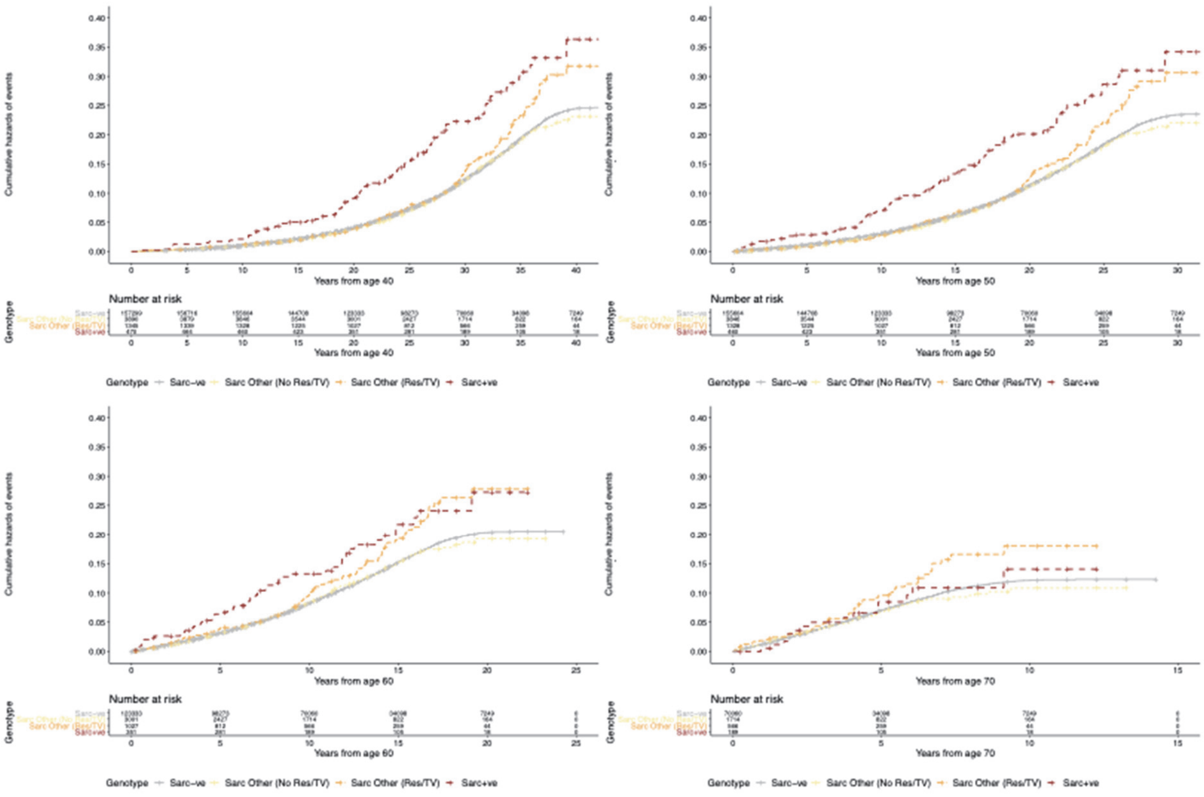

Cumulative hazard curves of landmark analysis for death or MACE stratified by genotype with SARC-IND separated into those with high probability and low probability variants. (Top panel left-right) from age 40, from age 50, (bottom panel left-right) from age 60 and from age 70. MACE defined as heart failure, arrhythmia, stroke and cardiac arrest events.
